# Systems vaccinology identifies clinical and immunological correlates of SARS-CoV-2 vaccine response in solid-organ transplant recipients

**DOI:** 10.1101/2024.04.05.24305357

**Authors:** Nicolas Gemander, Julika Neumann, Rafael Veiga, Isabelle Etienne, Teresa Prezzemolo, Delphine Kemlin, Pieter Pannus, Stéphanie Depickère, Véronique Olislagers, Inès Vu Duc, Alexandra Waegemans, Margaux Gerbaux, Leoni Bücken, Hafid Dahma, Charlotte Martin, Nicolas Dauby, Maria E Goossens, Isabelle Desombere, Carlos P Roca, Mathijs Willemsen, Stanislas Goriely, Alain Le Moine, Arnaud Marchant, Adrian Liston, Stephanie Humblet-Baron

## Abstract

Solid organ transplant (SOT) recipients are at enhanced risk of adverse outcomes following infectious challenges due to immunosuppressive treatment and additional comorbidities. Unfortunately, SOT recipients are also poor responders to the key medical intervention to preventing infection: vaccines. Here we performed a systems vaccinology study on a cohort of 59 kidney transplant recipients and 31 lung transplant recipients who received the mRNA Pfizer-BioNTech COVID-19 vaccine. Analyzing the immunological status of the patients prior to vaccination, we were able to identify multiple immunological associates of relatively improved vaccine responses following two or three doses of mRNA-based SARS-CoV-2 vaccine. These immunological associates predicted, with 95.0% and 93.3% accuracy, vaccine response after the second and third dose, respectively. Comparison of the immunological associates with vaccine response in SOT recipients revealed two distinct immune configurations: a non-classical configuration, distinct from the immune state of healthy subjects, associated with responses to two doses of mRNA vaccine and that could be mediated partly by the presence of double negative B cell subsets which are more prominently represented in responsive SOT recipients, and a “normalized” configuration, closer to the immune state of healthy subjects, associated with potent antibody responses to three doses of mRNA vaccine. These results suggest that immunosuppression in SOT recipients can result in distinct immune states associated with different trade-offs in vaccine responsiveness. Immune phenotyping of SOT recipients for immune constellation may be an effective approach for identifying patients most at risk of poor vaccine responses and susceptibility to vaccine-preventable diseases.

**One-sentence summary:** SOT recipients showed distinct immune states at baseline associated with different profiles of vaccine-associated immune response.

## Introduction

The recent pandemic, caused by the SARS-CoV-2 virus, resulted in more than six million deaths worldwide^1^ with a mortality rate about 2.3% in the general population^2^ as reported in early February 2020. Solid organ transplant (SOT) recipients were at higher risk of severe COVID-19 and death^3–8^ due to immunosuppressive therapies and frequent comorbidities, including cardiovascular disease, diabetes, and obesity.^6,9,10^ This higher susceptibility to infection by other pathogens had been previously described and related to predisposing factors such as diabetes^11^, renal dysfunction^12^, surgery^13^, nutritional deficits^14^ or modifications of gut microbiota^15^ in addition to immunosuppressive treatment regimens.^16,17^ Immune system alterations in SOT recipients, notably related to their immunosuppressive treatments^17^, are associated with lower vaccine responses.^18–20^ As SOT recipients had impaired humoral and cellular immune responses to primary COVID-19 mRNA vaccination compared to healthy adults^21–23^, these patients were prioritized for booster doses of COVID-19 vaccine.^24,25^

Despite vaccines being one of the fundamental tools in preventative medicine, the immunological correlates of effective vaccine responses remained largely unknown. Systems immunology has emerged as an indispensable tool to study the immune response in-depth in clinical settings. Using multi-parameter data from high-throughput sources, systems immunology was able to unravel and predict the immune response under specific perturbation. Specifically, in systems vaccinology studies, key components of the cellular adaptive immune system such as increased B cell activity with high plasmablasts within a week after vaccination have been correlated with protective antibody titers in healthy individuals.^26,27^ Importantly, this approach has highlighted the significant role of the innate immune compartment in vaccine response which had been disregarded until then, where early type I interferon (IFN) response promotes protective antibody response after vaccination, specifically after receiving the influenza vaccine.^26,27^ Interestingly, recent studies have integrated and compared the immune response of up to 13 different vaccines and, with the exception of few examples such as yellow fever, most of the vaccine responses elicit similar patterns with both higher adaptive immune response after one week and activation of the innate immune system already present at day 1 post-vaccination.^28,29^ Similar studies have been conducted with COVID-19 vaccines showing innate and adaptive immune responses after vaccination, the formet notably enhanced after the second immunization as compared to primary vaccination with BNT162b2 mRNA vaccine.^30,31^ Further, using a systems vaccinology approach, CD38^+^ helper T cell subsets induced soon after the second vaccine dose were identified as a prediction marker of neutralizing antibody titers 1 month after the second vaccination.^31^ These studies have been mainly performed in healthy adults and the degree to which their results will hold true in particular at-risk patient subsets, such as SOT recipients, remains largely unknown.

In addition, the moderate efficiency of COVID-19 vaccination in SOT recipients and the paucity of data on the immune configuration at baseline in this immunocompromised population support the need for dedicated systems vaccinology studies. Here, we used a systems vaccinology approach to identify the correlates of vaccine response in a cohort of 31 lung transplant and 59 kidney transplant recipients recruited at the beginning of the COVID-19 vaccination campaign. Patients were vaccinated with Pfizer-BioNTech COVID-19 mRNA vaccine Comirnaty, and both linear regression and machine learning approaches were used to identify demographic, medical and baseline immunological correlates of effective antibody response to mRNA vaccination. The results identify demographic and clinical features of the most at-risk patients for vaccine failure and highlight the negative impact of mycophenolate mofetil (MMF) and more recent immunosuppression induction therapy on the immune response to mRNA vaccination. Our results further suggest that vaccine responsiveness of SOT recipients can be achieved through either a non-traditional immune constellation at baseline capable of early but suboptimal responses, or via classical immune pathways requiring a third dose but leading to higher antibody titers more likely to confer some degree of protection against SARS-CoV-2.

## Results

### Systems vaccinology cohort design for identifying correlates with vaccine response in SOT recipients

To identify the pre-vaccination correlates for vaccine response in an immunosuppressed transplantation cohort, 90 patients were recruited to a systems vaccinology study. The study population included 31 lung and 59 kidney transplant recipients. Sex representation within these cohorts was 67.7% and 55.9% male, respectively and the median age of participants was 59 and 63 years. The median time post-transplantation was 5 years in the lung transplant group and 9.6 years in the kidney transplant group. SOT recipients were on different combinations of immunosuppressive treatments during vaccination, including corticosteroids (CS), antimetabolites such as azathioprine (AZA) and mycophenolate mofetil, calcineurin inhibitors such as tacrolimus (FK506) and cyclosporin A (CsA), and the mTOR inhibitor everolimus. Full demographic breakdown of the study cohort is shown in **Table 1**. Patients were vaccinated with the Pfizer-BioNTech COVID-19 vaccine Comirnaty and were categorized exhibiting no response (<5 binding antibody units (BAU)/mL), detectable response (detectable threshold at >5 BAU/mL) or positive response (positive threshold at >264 BAU/mL as used in previous studies^32–34)^.

**Table 1:** Demographics of study population. SOT: solid organ transplant, HTA: hypertension, CVD: cardiovascular disease, eGFR: estimated glomerular filtration rate, CS: Corticosteroids, AZA: Azathioprine, MMF: Mycophenolate mofetil, FK506: Tacrolimus, CsA: Cyclosporin A, Evero: Everolimus, ATG: anti-thymocyte globulin, OKT3: anti-CD3. ---: data was not available, NA: not applicable. Data on induction treatment was missing for two lung and two kidney transplant recipients. One lung transplant recipient did not receive any induction treatment. * Chi-squared test. Significant comparisons are highlighted in bold.

| Variables | Healthy controls<br>(n= 20 \| 18.2%) |  | Lung transplant<br>(n= 31 \| 28.2%) |  | Kidney transplant<br>(n= 59 \| 53.6%) |  | Lung vs<br>Kidney | Total SOT<br>(N= 90) |  |
| --- | --- | --- | --- | --- | --- | --- | --- | --- | --- |
| General | n | % | n | % | n | % | p-value* | n | % |
| Sex (male) | 13 | 65.0 | 21 | 67.7 | 33 | 55.9 | 0.389 | 54 | 60.0 |
| Age |  |  |  |  |  |  | **0.039** |  |  |
| <50 | 0 | 0.0 | 12 | 38.7 | 11 | 18.6 |  | 23 | 25.6 |
| 50-70 | 20 | 100.0 | 18 | 58.1 | 38 | 64.4 |  | 56 | 62.2 |
| >70 | 0 | 0.0 | 1 | 3.2 | 10 | 16.9 |  | 11 | 12.2 |
| BMI |  |  |  |  |  |  | 0.183 |  |  |
| Standard | --- | --- | 19 | 61.3 | 26 | 44.1 |  | 45 | 50.0 |
| Overweight | --- | --- | 12 | 38.7 | 33 | 55.9 |  | 45 | 50.0 |
| Ethnicity |  |  |  |  |  |  | 0.423 |  |  |
| European | --- | --- | 28 | 90.3 | 48 | 81.4 |  | 76 | 84.4 |
| African | --- | --- | 3 | 9.7 | 9 | 15.3 |  | 12 | 13.3 |
| Other | --- | --- | 0 | 0.0 | 2 | 3.4 |  | 2 | 2.2 |
| Time after transplant |  |  |  |  |  |  | **<0.001** |  |  |
| >10 years | NA | NA | 9 | 29.0 | 28 | 47.5 |  | 37 | 41.1 |
| 3-10 years | NA | NA | 14 | 45.2 | 18 | 30.5 |  | 32 | 35.6 |
| <3 years | NA | NA | 8 | 25.8 | 13 | 22.0 |  | 21 | 23.3 |
| Transplant rank |  |  |  |  |  |  | 0.450 |  |  |
| One | NA | NA | 30 | 96.8 | 53 | 89.8 |  | 83 | 92.2 |
| Two or more | NA | NA | 1 | 3.2 | 6 | 10.2 |  | 7 | 7.8 |
| **Comorbidities** |  |  |  |  |  |  |  |  |  |
| Diabetes | 0 | 0.0 | 13 | 41.9 | 22 | 37.3 | 0.840 | 35 | 38.9 |
| HTA | 5 | 25.0 | 20 | 64.5 | 49 | 83.1 | 0.087 | 69 | 67.7 |
| CVD | 0 | 0.0 | 2 | 6.5 | 16 | 27.1 | **0.040** | 18 | 20.0 |
| eGFR <30 | 0 | 0.0 | 2 | 6.5 | 8 | 13.6 | 0.505 | 10 | 11.1 |
| Non-invasive cancer | 1 | 5.0 | 1 | 3.2 | 15 | 25.4 | **0.020** | 16 | 17.8 |
| **Treatment** |  |  |  |  |  |  |  |  |  |
| CS | NA | NA | 31 | 100.0 | 46 | 78.0 | **0.012** | 77 | 85.6 |
| AZA | NA | NA | 4 | 12.9 | 15 | 25.4 | 0.266 | 19 | 21.1 |
| MMF | NA | NA | 16 | 51.6 | 30 | 50.8 | 0.999 | 46 | 51.1 |
| FK506 | NA | NA | 31 | 100.0 | 40 | 67.8 | **0.001** | 71 | 78.9 |
| CsA | NA | NA | 0 | 0.0 | 8 | 13.6 | 0.079 | 8 | 8.9 |
| Evero | NA | NA | 0 | 0.0 | 14 | 23.7 | **0.008** | 14 | 15.6 |
| Induction |  |  |  |  |  |  | **<0.001** |  |  |
| ATG | NA | NA | 28 | 100.0 | 18 | 31.6 |  | 46 | 54.1 |
| Anti-IL2R | NA | NA | 0 | 0.0 | 34 | 59.6 |  | 34 | 40.0 |
| OKT3 | NA | NA | 0 | 0.0 | 5 | 8.8 |  | 5 | 5.9 |
| Number of immunosuppressive treatments |  |  |  |  |  |  | 0.800 |  |  |
| Two | NA | NA | 11 | 35.5 | 24 | 40.7 |  | 35 | 38.9 |
| Three | NA | NA | 20 | 64.5 | 35 | 59.3 |  | 55 | 61.1 |

Overall, vaccine response rates were low, as previously reported.^22,24^ The transplantation cohort had 0% of positive responses after a single dose (assessed at day 21, before D2), increasing to 3.4% after two doses (assessed at day 49, 28 days after D2) and 24.0% after three doses (assessed at day ∼188, 28 days after D3). Detectable responses were, however, observed in 6.6%, 39.8% and 49.4% of the transplantation cohort, after 1, 2 and 3 doses of the vaccine, respectively (**Supplementary Figure 2**). This study design allowed for the determination of demographic, clinical and baseline (pre-vaccination) immunological parameters that correlate with detectable or positive responses, after 2 or 3 doses of COVID-19 mRNA vaccine, respectively. As BNT162b2 vaccine has shown >90% efficacy in preventing COVID-19 illness in the general population^35^, twenty healthy controls were included in our study for comparing their classical immune profile at baseline to that of SOT recipients. Their clinical characteristics are summarized in **Table 1**.

### Demographic and clinical correlates for vaccine response in SOT recipients

We first investigated the effect of demographic and clinical variables on the vaccine response. Using a linear regression approach, no significant difference was observed between kidney and lung transplant recipients when assessing detectable vaccine responses after 2 or 3 doses (**Table 2**). However, a smaller proportion of lung transplant recipients showed a positive response after dose 3 compared to kidney transplant recipients. Within all the SOT recipients, no significant effect was observed for BMI or ethnicity. Although not significant, a sex effect was observed, with males more likely to have a positive vaccine response after dose 3. A complex relationship was observed with age – superior vaccine responses were observed in patients aged <50, while vaccine responses were significantly reduced in patients aged 50-70, consistent with a known association of age with reduced vaccine efficacy.^36^ This effect was however blunted in the oldest patient cohort, aged between 71 and 90 years old, an observation that can potentially be explained by a longer period since transplantation within this group (**Supplementary Figure 3A**) especially given that years post-transplantation were positively associated with vaccine response (with poorest responses in those <3 years post-transplantation and significant improvements in those >10 years post-transplantation). Moreover, the reduced effect of age on vaccine response in this oldest patient cohort could be associated with a reduced number of cumulative immunosuppressive treatments impacting the immune response too (**Supplementary Figure 3B**), as well as a selection bias for survivors of infections suggesting a stronger residual immune response in this older group. We were also able to associate comorbidities of SOT recipients with vaccine response, with the only significant association being a detrimental effect of a estimated glomerular filtration rate (eGFR) <30 mL/min/1.73m^2^ after dose 2 as already described in the literature.^37^ In chronic kidney disease, uremia has notably been related to T cell defects both by decreasing thymic output and increasing T cell senescence.^38^ In addition, vaccine-induced immune response was impacted by immunosuppressive treatments, particularly in case of combination of multiple treatments for positive response after dose 3. However, as these drugs are ubiquitous within the poorly- responding transplant cohort, effects could only be assessed in comparison to competing treatment regimens. Using this comparison, MMF treatment was the only significant influence over vaccine response, with significantly poorer outcomes after each dose and using both the detectable and positive thresholds.

**Table 2:** Association between demographic variables, comorbidities, and treatment with vaccine response in SOT recipients. *Adjusted for sex, age, MMF, eGFR<30 and Induction treatment **Adjusted for sex, age, MMF, time after transplant and eGFR<30. ***Adjusted for sex, age, MMF, eGFR<30, transplant type, number of treatments, CsA and time after transplant. ****not adjusted for the number of treatments. ***** not adjusted for any other treatment. Statistically significant values are in bold. Detectable vaccine response was defined as >5 BAU/mL and positive response as >264 BAU/mL. OR: odds ratio, HTA: hypertension, CVD: cardiovascular disease, eGFR: estimated glomerular filtration rate, CS: Corticosteroids, AZA: Azathioprine, MMF: Mycophenolate mofetil, FK506: Tacrolimus, CsA: Cyclosporin A, Evero: Everolimus, ATG: anti-thymocyte globulin, OKT3: anti-CD3.

| General Variables | Detectable for d2<br>(n=35 \| 39.8%)* |  | Detectable for d3<br>(n=39 \| 49.4%)** |  | Positive for d3<br>(n=19 \| 24.0%*** |  |
| --- | --- | --- | --- | --- | --- | --- |
|  | n (% within variable group) | OR (95% C.I.) | n (% within variable group) | OR (95% C.I.) | n (% within variable group) | OR (95% C.I.) |
| Sex (male) | 19(36.5) | 1.18(0.39-3.74) | 25(50.0) | 1.22(0.39-3.89) | 13(26.0) | 1.55(0.32-8.62) |
| Age |  |  |  |  |  |  |
| <50 | 8(34.8) | 1 | 13(68.4) | 1 | 7(36.8) | 1 |
| 50-70 | 22(40.7) | 0.66(0.15-2.64) | 21(40.4) | 0.12(0.02-0.48) | 9(17.3) | 0.03(0.00-0.22) |
| >70 | 5(45.5) | 2.54(0.29-27.57) | 5(62.5) | 0.61(0.05-15.70) | 3(37.5) | 0.67(0.02-50.05) |
| BMI |  |  |  |  |  |  |
| Standard | 15(34.1) | 1 | 21(51.2) | 1 | 9(22.0) | 1 |
| Overweight | 20(45.5) | 1.80(0.62-5.47) | 18(47.4) | 0.77(0.25-2.29) | 10(26.3) | 0.92(0.21-3.93) |
| Ethnicity |  |  |  |  |  |  |
| European | 29(39.2) | 1 | 32(47.8) | 1 | 13(19.4) | 1 |
| African | 6(50.0) | 1.34(0.27-6.93) | 6(54.5) | 1.49(0.24-10.23) | 5(45.5) | 0.51(0.04-4.63) |
| Other | 0(0.0) | --- | 1(100.0) | --- | 1(100.0) | --- |
| Transplant type |  |  |  |  |  |  |
| Kidney | 24(42.1) | 1 | 25(50.0) | 1 | 16(32.0) | 1 |
| Lung | 11(35.5) | 0.71(0.17-3.01) | 14(48.3) | 0.73(0.22-2.27) | 3(10.3) | 0.10(0.02-0.44) |
| Time after transplant |  |  |  |  |  |  |
| >10 years | 17(47.2) | 1 | 19(61.3) | 1 | 11(35.5) | 1 |
| 3-10 years | 13(41.9) | 1.01(0.28-3.75) | 16(55.2) | 0.96(0.28-3.29) | 6(20.7) | 0.82(0.15-4.86) |
| <3 years | 5(23.8) | 0.34(0.07-1.47) | 4(21.1) | 0.17(0.03-0.70) | 2(10.5) | 0.13(0.01-1.11) |
| Transplant rank |  |  |  |  |  |  |
| One | 34(42.0) | 1 | 37(50.0) | 1 | 18(24.3) | 1 |
| Two or more | 1(14.3) | 0.34(0.02-3.19) | 2(40.0) | 0.86(0.07-15.30) | 1(20.0) | 1.70(0.04-79.43) |
| **Comorbidities** |  |  |  |  |  |  |
| Diabetes | 16(47.1) | 1.50(0.52-.42) | 18(54.5) | 2.08(0.69-6.72) | 11(33.3) | 1.86(0.44-8.15) |
| HTA | 27(39.7) | 0.53(0.14-1.98) | 29(48.3) | 0.92(0.23-3.54) | 14(23.3) | 0.50(0.09-2.77) |
| CVD | 6(33.3) | 0.54(0.11-2.33) | 5(29.4) | 0.64(0.13-2.93) | 4(23.5) | 0.58(0.07-3.81) |
| eGFR<30 | 1(10.0) | 0.03(0.00-0.33) | 2(28.6) | 0.13(0.00-1.58) | 0(0.0) | --- |
| Cancer | 8(50.0) | 0.80(0.17-3.64) | 6(50.0) | 0.49(0.09-2.47) | 4(33.3) | 1.02(0.12-8.37) |
| **Treatment\*\*\*\*** |  |  |  |  |  |  |
| CS | 31(41.3) | 0.98(0.20-5.11) | 34(50.0) | 0.59(0.08-4.01) | 15(22.1) | 0.11(0.00-1.47) |
| AZA | 13(68.4) | 2.31(0.48-13.35) | 13(72.2) | 1.86(0.36-10.06) | 6(33.3) | 0.45(0.06-2.90) |
| MMF | 10(22.2) | 0.12(0.03-0.39) | 16(40.0) | 0.18(0.05-0.57) | 7(17.5) | 0.12(0.01-0.55) |
| FK506 | 28(40.6) | 1.85(0.44-8.86) | 32(52.5) | 1.90(0.52-7.41) | 15(24.6) | 0.96(0.15-6.63) |
| CsA | 3(37.5) | 0.62(0.05-4.92) | 3(37.5) | 0.30(0.04-1.89) | 1(12.5) | 0.08(0.00-1.07) |
| Evero | 6(46.2) | 0.97(0.20-4.73) | 7(50.0) | 1.57(0.36-7.24) | 4(28.6) | 0.77(0.11-4.84) |
| Induction |  |  |  |  |  |  |
| ATG | 17(37.8) | 1 | 20(45.5) | 1 | 7(15.9) | 1 |
| Anti-IL2R | 13(39.4) | 1.39(0.46-4.32) | 14(51.9) | 1.13(0.33-3.98) | 10(37.0) | 1.42(0.28-7.69) |
| OKT3 | 2(40.0) | 0.99(0.09-11.69) | 2(50.0) | 0.86(0.07-11.90) | 0(0.0) | --- |
| Number of immunosuppressive treatments***** |  |  |  |  |  |  |
| Two | 14(40.0) | 1 | 12(42.9) | 1 | 9(32.1) | 1 |
| Three | 21(39.6) | 1.24(0.46-3.52) | 27(52.9) | 1.16(0.35-3.92) | 10(19.6) | 0.21(0.04-0.94) |

### Multiple immune configurations associated with vaccine response in SOT recipients

We next investigated the effect of the baseline immunological profile (immune profile at day 0 of vaccination) on subsequent antibody production. Blood samples from each patient were assessed by flow cytometry for 444 immunological variables, constituting a deep phenotypic analysis of the adaptive immune compartment with extensive innate immune profiling. Two major divisions of the transplant recipients cohort were used: patients capable of some response after two doses of vaccine (“detectable” after second dose versus “undetectable” after second dose), and patients capable of a strong antibody response (>264 BAU/mL) after three doses of vaccine (“positive” after third dose versus “not-positive” after third dose). Of note, these 2 categories represent two different immune profiles at baseline associated with immune response after 2 and 3 doses of mRNA vaccine. However, the majority of patients showing a positive response after dose 3 had a detectable response after dose 2 implying an overlap of patients between these 2 baseline profiles. Multivariate logistic regression was performed adjusting by type of transplant, sex, age and the number of years since transplantation, to identify the immunological associates corresponding to vaccine response. Of the immunological parameters associated with vaccine response after demographic adjustment (**Supplementary Spreadsheet 3**), the 10 parameters with greatest magnitude effects were further analyzed (**Figure 1**). For some response (“detectable”) after dose 2, the strongest positively associated parameter was CD80 expression, with CD80+ median expression on double negative CD27^-^IgD^-^ B cells DN1 (CD11c^-^CD21^+^) and CD21^-^ B cells including CD21^-^CD27^+^ B cells along with the frequency of CD80+ double negative B cells DN2 (CD11c^+^CD21^-^) (**Figure 1A**). For strong vaccine responses (“positive” after dose 3), the key immunological parameters differed: the strongest positive associations were with non-classical monocytes and CD40^+^ plasmacytoid dendritic cells, within their respective parent populations, along with CD40^+^ median expression in CD21^-^ B cells and double negative B cells DN2 (**Figure 1B**). In both cases, the immunological dataset was from baseline (day 0) data and showed associations of different immunological parameters between some response after dose 2 and a strong response after dose 3. These associations highlight the different baseline profiles associated with different outcomes. One of the main differences was the baseline T cell activation that was rather predictive of a negative response after dose 3. Thus, the immunological profiles associated with a detectable vaccine response after a two-dose mRNA vaccine schedule were distinct from the immunological profiles associated with a positive vaccine response after a third injection of mRNA vaccine (booster) among SOT recipients.

**Figure 1.**
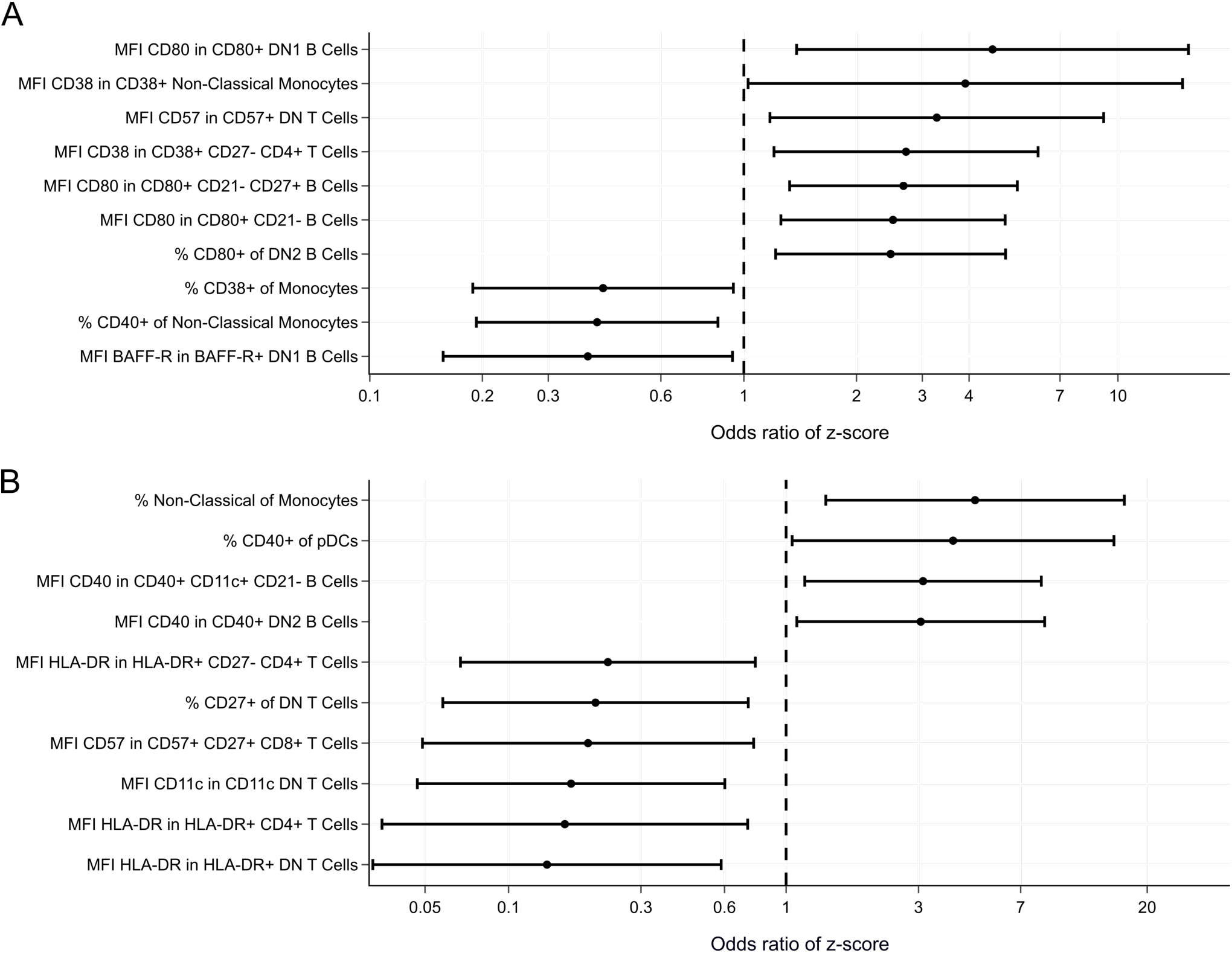
Identification of key immune parameters associated with effective SARS-CoV-2 vaccine response. 110 individuals were recruited to a systems vaccinology study, including 20 healthy controls, 31 lung transplant recipients and 59 kidney transplant recipients. Immunological profiles were assessed at baseline (day 0 of vaccination) for 444 immunological variables. Multivariate logistic regression was performed adjusting by type of transplant, sex, age and the number of years since transplantation, to identify the immunological associates with vaccine response. **A.** Odds ratio (OR) of the z-score of the 10 highest difference parameters (magnitude cut-off), using the stratification of transplant recipients “detectable” or “undetectable” for SARS-CoV-2 antibodies after two doses of the mRNA vaccine (day 49). **B.** Odds ratio (OR) of the z-score of the 10 highest difference parameters (magnitude cut-off), using the stratification of transplant recipients “positive” or “non-positive” for SARS-CoV-2 antibodies after three doses of the mRNA vaccine (day ∼188).

In identifying immunological parameters associated with effective vaccine response after the second and the third dose, our *a priori* hypothesis was that the responding phenotype, as opposed to the non-responding phenotype, of SOT recipients would be more similar to the immune status at baseline of healthy subjects. For each of the main immune associates, we compared the values of both responder and non-responder SOT recipients to those of healthy controls. For the ability to have some response after mRNA vaccination (dose 2 “detectable”), of the 10 strongest immune associates in SOT recipients, surprisingly 7 showed a clear trend of the non-detectable SOT population being more similar to the healthy controls, while 1 showed the expected pattern of the detectable SOT cohort being more similar to the healthy controls (**Figure 2A**). These immune parameters did not seem to be directly impacted by the immunosuppressive treatment regimen. The strongest suppression of vaccine responses was associated with MMF and CS treatment (**Table 2**), yet segregation of “detectable” and “non-detectable” patients by MMF (**Supplementary Figure 4A**) or CS (**Supplementary Figure 5A**) found few consistent patterns. One exception was the frequency of CD38+ monocytes, a rare case where the “detectable” SOT population had a phenotype closer to that of healthy controls. This specific association was driven instead by CS treatment, and assessing only the CS untreated patients, the “detectable” population was actually further away from the healthy control population (**Supplementary Figure 5A**).

**Figure 2.**
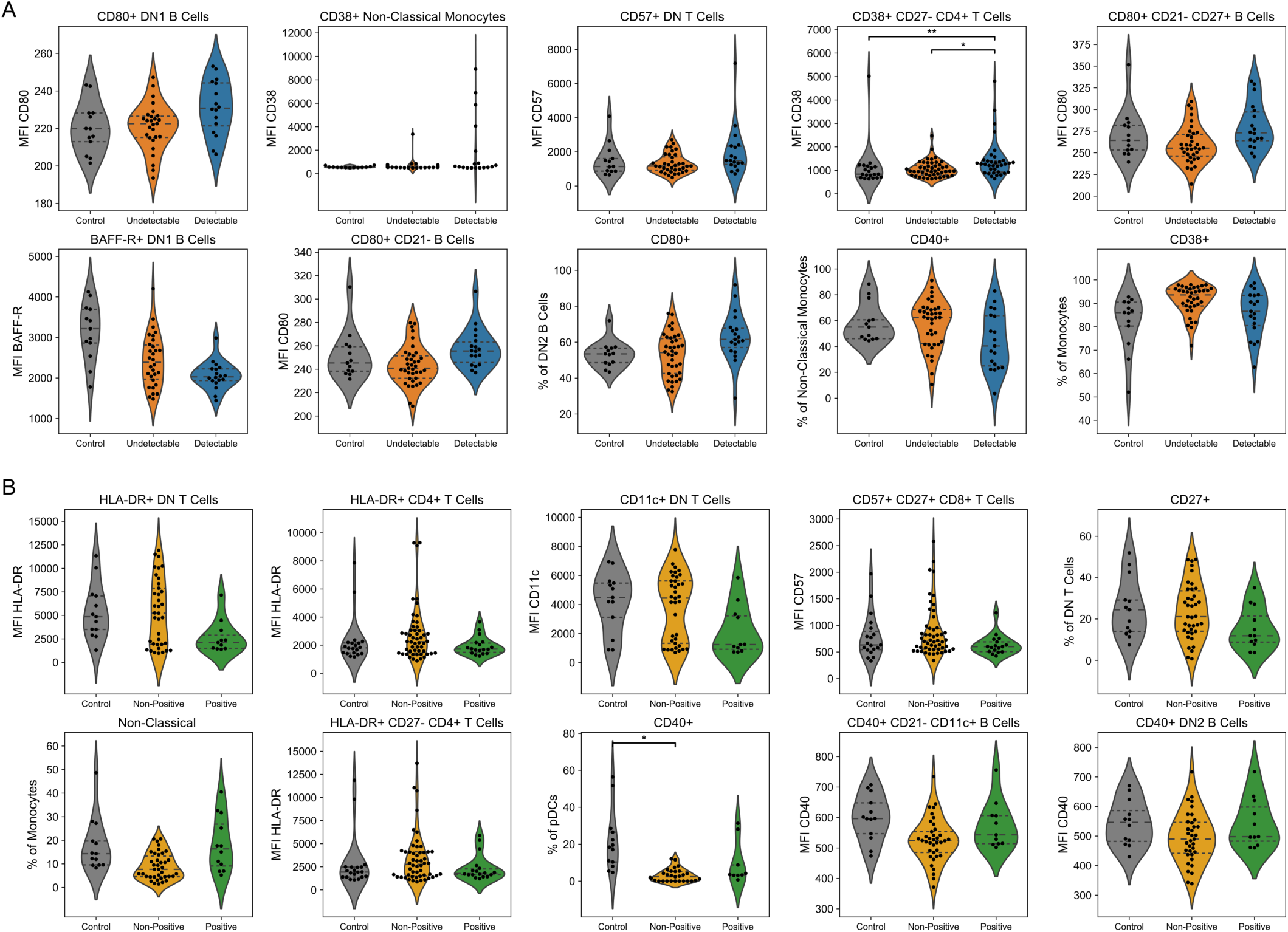
Key immune changes associated with effective SARS-CoV-2 vaccine response in SOT recipients. 110 individuals were recruited to a systems vaccinology study, including 20 healthy controls, 31 lung transplant recipients and 59 kidney transplant recipients. Immunological profiles were assessed at baseline (day 0 of vaccination) for 444 immunological variables. Multivariate logistic regression was performed adjusting by type of transplant, sex, age and the number of years since transplantation, identifying the 10 highest magnitude effects (based on log(OR of the z-score)). **A.** Raw value graphs, showing healthy control, dose 2 non- detectable and dose 2 detectable SOT recipients, for the 10 highest magnitude parameters associated with dose 2 antibody detection. Individual values, no imputation. **B.** Raw value graphs, showing healthy controls, dose 3 non-positive and dose 3 positive SOT recipients, for the 10 highest magnitude parameters associated with dose 3 antibody positive seroconversion. Individual values, no imputation. Statistics are based on Kruskal-Wallis with Dunn’s post-hoc comparison and only significant comparisons are shown. * = p<0.05; ** = p<0.01.

In contrast to the analysis for some ability to respond, for the ability to have a strong response after a booster dose of mRNA vaccine (“positive” after dose 3), most of the parameters showed a clear trend for the positive SOT population being more similar to the healthy controls, while parameters on double negative CD4^-^CD8^-^ T cells that had the non-positive SOT population were more similar to the healthy controls (**Figure 2B**). Again, these immune parameters were largely not directly impacted by treatment regimen (**Supplementary Figure 4B, Supplementary Figure 5B**). These results suggest two distinct functional immune profiles in SOT recipients: a specific non-classical immune profile, capable to develop a detectable humoral response after 2 doses of mRNA vaccine, and a distinct immune profile closer to the healthy population, which is related to a better response to vaccination after a third dose. The non-classical immune pattern might be driven by “unconventional” immune cell subsets, such as double negative (DN) B cells, a cellular subset associated with antibody-mediated response which is less abundant in the general population but has been involved in autoimmune diseases and in vaccine response.^39,40^ In our cohort, all responders displayed a higher amount of these DN B cells (**Supplementary Figure 6A**). In contrast, an immune profile of SOT recipients closer to the one found in the healthy population was able to elicit higher antibody levels after additional vaccine administration (dose 3). Recently, identification of an adaptive immune signature at baseline prior to vaccination in a healthy population was associated with a weaker antibody response^29^, and interestingly, our data show that T cell activation (such as HLA-DR expression on CD4^+^ T cells) reduced the chance to develop high antibody levels after the third vaccine dose (**Figure 2B**).

To assess this immunological configuration model, we performed two analyses of the global immune profile. First, we used a Uniform Manifold Approximation and Projection (UMAP) approach of the baseline immunological profile of SOT recipients, responders and non-responders, and healthy controls; considering the baseline immune profile of this latter group associated with a high immune response to 2 doses of mRNA vaccination as reported previously.^35^ For some level of response (“detectable” after dose 2), a slight separation was observed between “detectable” and healthy controls when displaying all 444 immune parameters; a stronger separation when only the 10 most highly associated immune parameters were displayed (**Figure 3A**). By contrast, when assessing the immunological profile of strong responders (“positive” after dose 3), the responders clustered closer to the healthy controls (**Figure 3B**), consistent with the individual parameter assessment described above. Second, we looked at the relationship between immune parameters using Spearman’s correlations. This allowed us to look at the relationship of the key immune parameters in healthy controls, and whether that relationship was preserved or disrupted in responding or non-responding SOT recipients. For the top 10 associated immune parameters after dose 2, we observed strong correlations between different immune characteristics within the healthy controls, for example a strong co-correlation of the CD80-associated phenotypes, and a strong negative correlation between CD57 expression on double negative T cells and the CD80-associated phenotypes (**Figure 4A**). In the SOT recipients capable of some response (“detectable”) after dose 2, large disturbances to these correlations were observed, while SOT recipients that did not respond (“not detectable”) after dose 2 showed more similar correlations as the healthy population (**Figure 4A,B**). By contrast, the disruptions in co-correlations in the non-responding and responding SOT recipients groups after dose 3 were similar (**Figure 4C,D**). Together, these analyses support a model where some SOT recipients exhibited a non-classical immune profile, capable of sub-optimal response after 2 vaccine doses, while some SOT recipients showed a more “normalized” immune profile in certain immune subsets, allowing higher levels of antibodies if an additional vaccine dose was given.

**Figure 3.**
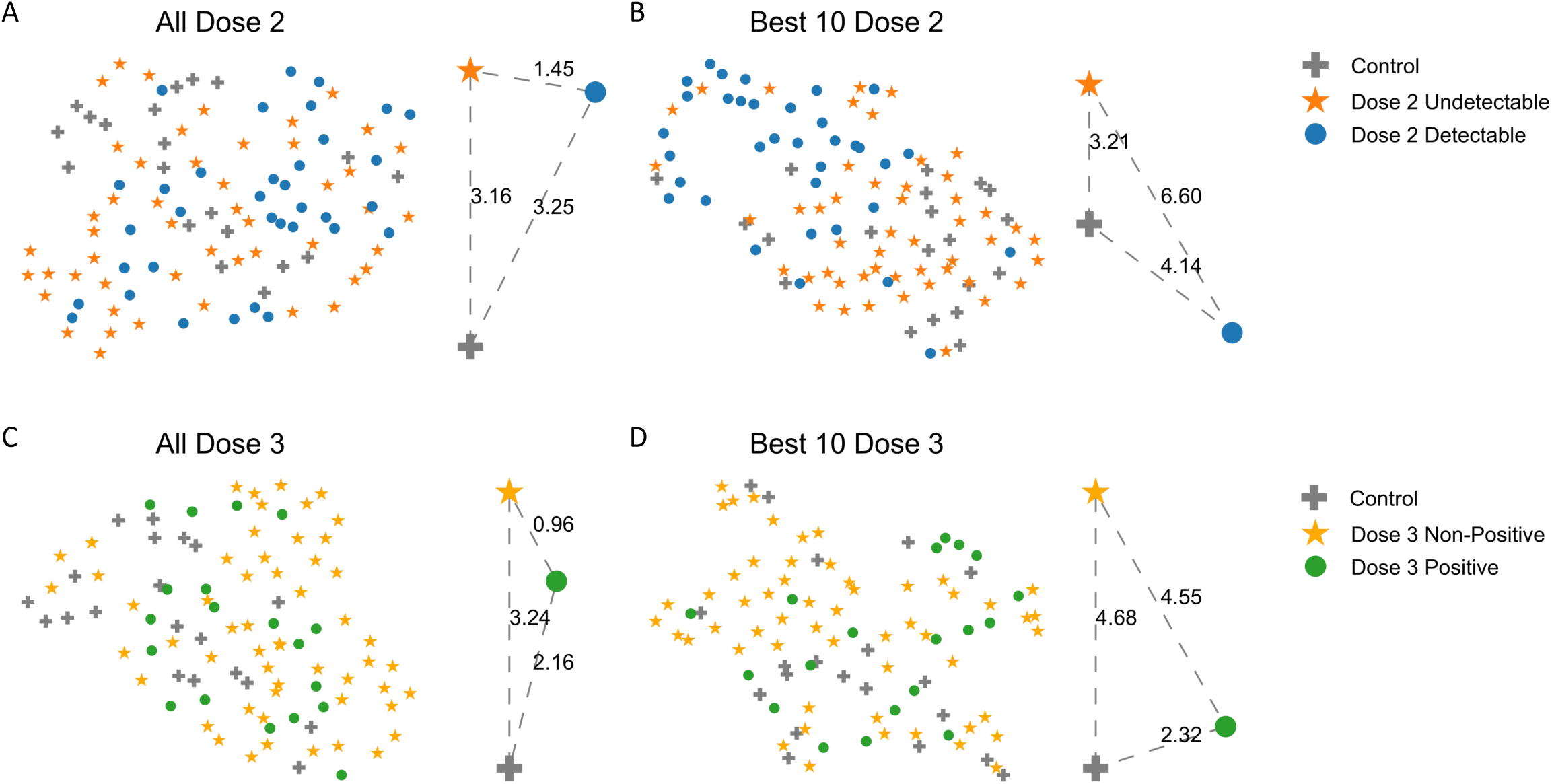
Immune configuration associated with effective SARS-CoV-2 vaccine response in SOT recipients. 110 individuals were recruited to a systems vaccinology study, including 20 healthy controls, 31 lung transplant recipients and 59 kidney transplant recipients. Immunological profiles were assessed at baseline (day 0 of vaccination) for 444 immunological variables, and SOT recipients were assessed for ability to generate binding antibodies after vaccination. Immune configurations displayed as UMAP plots for listed parameters for each individual, annotated based on patient characteristics. Distances between the groups were calculated using the Calinski and Harabasz score and are shown on the right of each panel. **A.** UMAP for all 444 all immune parameters, with individuals annotated as healthy controls, or SOT recipients with non-detectable or detectable antibodies after dose 2. **B.** UMAP for 10 highest magnitude parameters associated with “detectable” responses after dose 2, with the same patient annotation. **C.** UMAP for all 444 all immune parameters, with individuals annotated as healthy controls, or SOT recipients with non-positive or positive antibodies responses after dose 3. **D.** UMAP for 10 highest magnitude parameters associated with “positive” responses after dose 3, with the same patient annotation.

**Figure 4.**
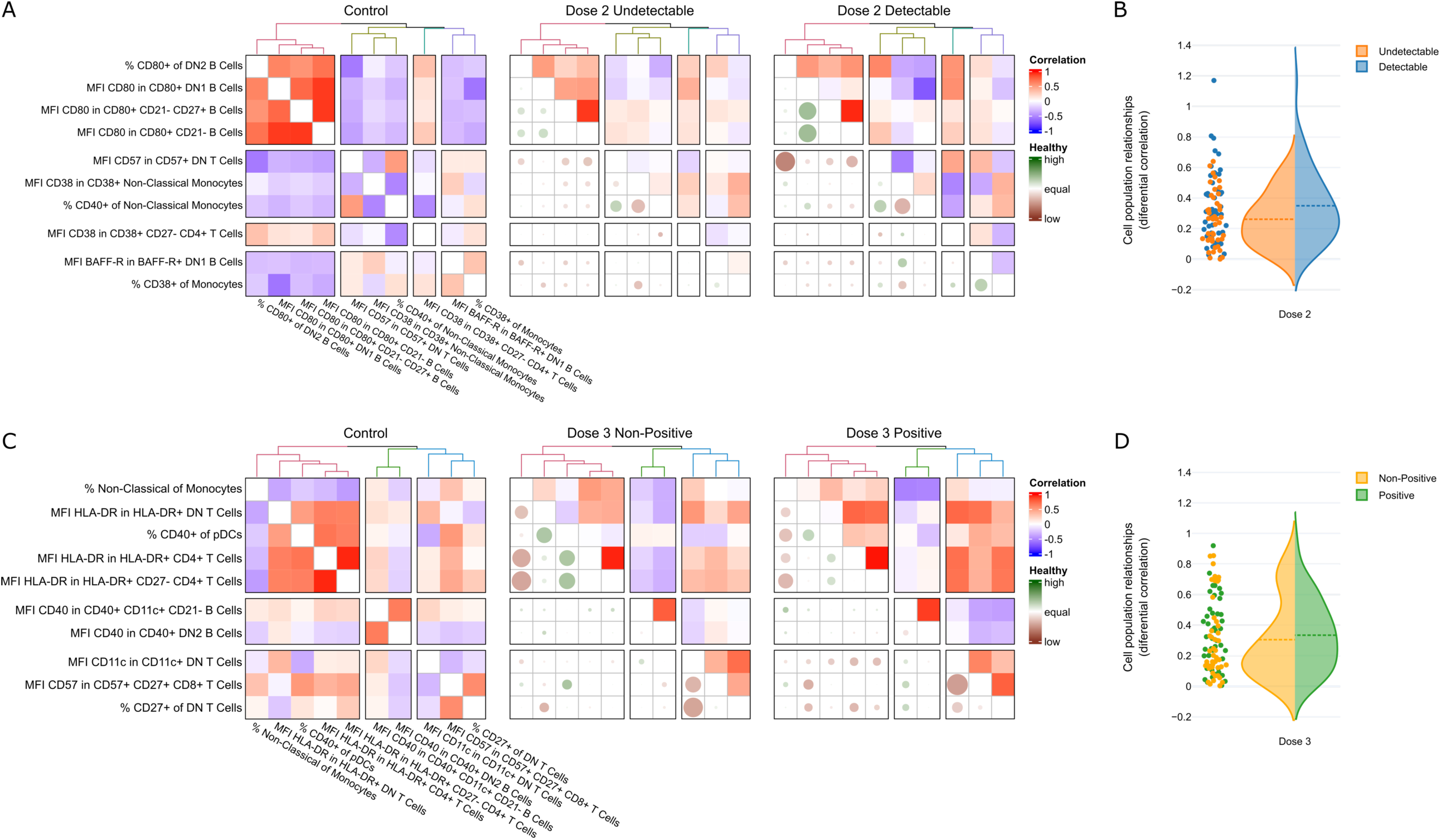
Immunological relationships associated with effective SARS-CoV-2 vaccine response in SOT recipients. 110 individuals were recruited to a systems vaccinology study, including 20 healthy controls, 31 lung transplant recipients and 59 kidney transplant recipients. Immunological profiles were assessed at baseline (day 0 of vaccination) for 444 immunological variables, and SOT recipients were assessed for ability to generate binding antibodies after vaccination. Multivariate logistic regression was performed adjusting by type of transplant, sex, age and the number of years since transplantation, identifying the 10 highest magnitude effects (based on log(OR of the z-score)). Spearman’s correlations were calculated between each of the 10 highest magnitude effect variables. **A.** For those immune parameters associated with dose 2 “detectable”, Spearman’s correlations between each parameter pair are shown for healthy controls (left). For dose 2 “undetectable” (center) and “detectable” (right) SOT recipients, the upper-right triangular matrix shows the Spearman’s correlations within the respective patient population, while the lower-left triangular matrix visualizes by color and size the deviations in the correlations observed between the patient population and the healthy population with green indicating high correlation within the healthy population and low correlation within the patient population and brown low correlation within the healthy population and high correlation within the patient population. **B.** Values of the differential correlation for each cell population associated with a detectable response after dose 2, comparing the healthy population and the respective patient population as individual values (left) and violin plot (right). **C.** For those immune parameters associated with dose 3 “positive”, Spearman’s correlations between each parameter pair are shown for healthy controls (left). For dose 3 “non-positive” (center) and “positive” (right) SOT recipients, the upper-right triangular matrix shows the Spearman’s correlations within the respective patient population, while the lower-left triangular matrix visualizes by color and size the deviations in the correlations observed between the patient population and the healthy population. **D.** Values of the differential correlation for each cell population associated with a positive response after dose 3, comparing the healthy population and the respective patient population as individual values (left) and violin plot (right).

### Machine learning identifies immunological parameters predicting vaccine response

Our results demonstrate the influence of baseline immune configuration on vaccine response in SOT recipients. To identify those immune parameters with the highest potential utility in diagnosis, we used a machine learning approach. Multiple machine learning models were used and compared by recursive feature elimination, with 10-fold cross-validation to assess the explanatory power of the model, with the area under the ROC curve (AUC). For both the dose 2 “detectable” and dose 3 “positive” classifications, the support vector machine approach gave the highest accuracy (**Table 3**). For some level of response (“detectable” after dose 2), the optimal machine learning model had a predictive capacity (AUC) of 95% (**Figure 5C**), while for strong responses (“positive” after dose 3), the optimal machine learning model had a predictive capacity (AUC) of 93.3% (**Figure 5D**). To identify the immunological parameters contributing to the predictive model, we used a cluster permutation approach. For some response (“detectable” after dose 2), the strongest single parameter was the frequency of CD40^+^ B cells, with NK and NKT phenotypes also strongly contributing, and CD80^+^ median expression in switched memory B cells as a strong influence (**Figure 5A**). For strong response (“positive” after dose 3), the strongest single parameter was CD40^+^ median expression in non-switched memory B cells, with CD40 expression in naive B cells also ranking highly, along with NK phenotypes (**Figure 5B**). Notably, while there is some overlap with the linear regression multivariate analysis used above, the lists of features are distinct, as highly co-correlated parameters (such as CD80 expression across multiple B cell subtypes) provide decreasing amounts of additional information, and therefore do not dominate the influential parameter list in machine learning approach. Together, this analysis suggests the key markers aiming to stratify SOT recipients into different functional groups regarding vaccine response outcome.

**Figure 5.**
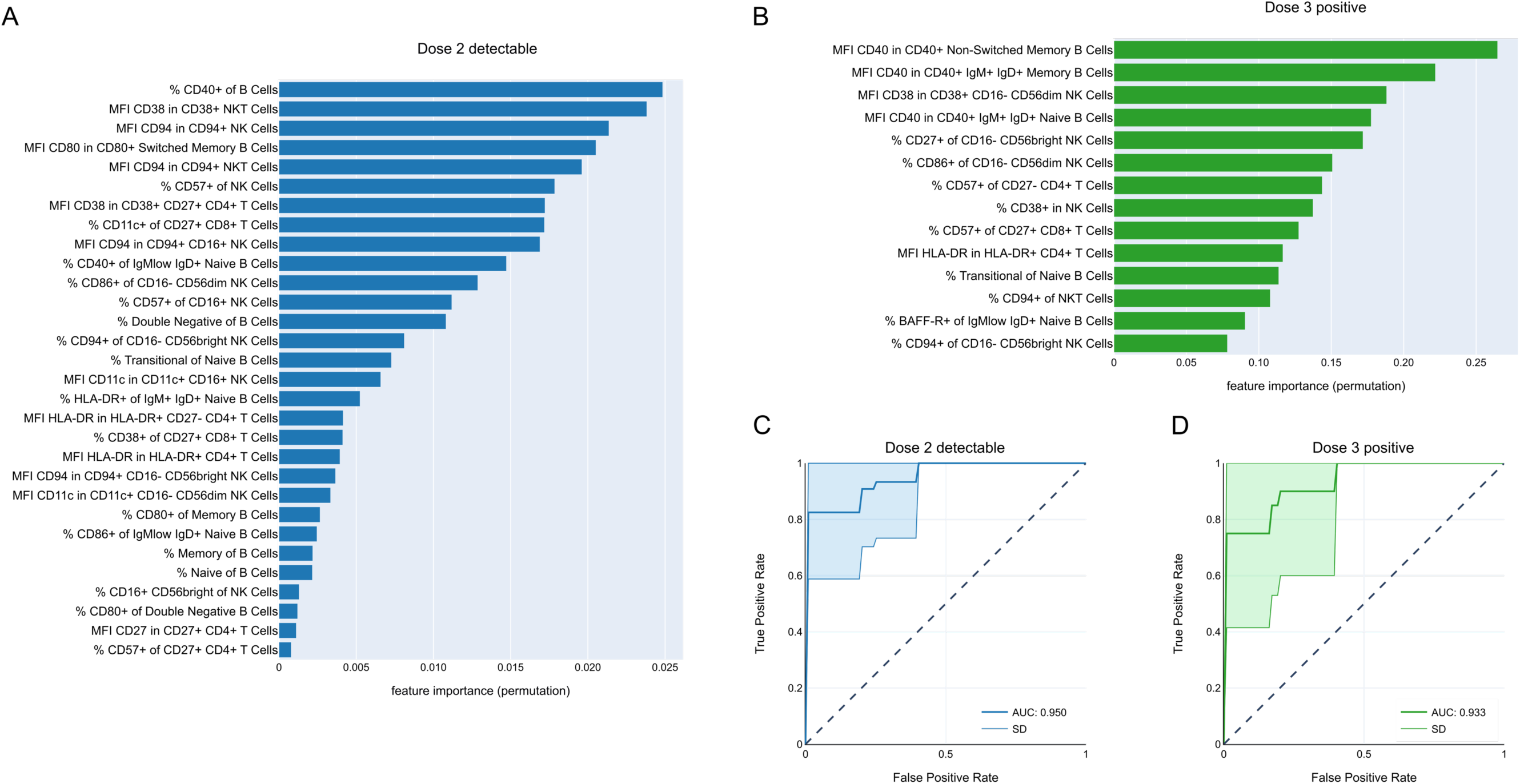
Machine learning predicts SARS-CoV-2 vaccine response in SOT recipients. 110 individuals were recruited to a systems vaccinology study, including 20 healthy controls, 31 lung transplant recipients and 59 kidney transplant recipients. Immunological profiles were assessed at baseline (day 0 of vaccination) for 444 immunological variables, and SOT recipients were assessed for ability to generate binding antibodies after vaccination. The support vector machine approach was selected to model vaccine response, based on the highest performance. **A.** Best features according to the machine learning model to identify the detection of antibodies against COVID-19 after the second dose of the vaccine. The best features were chosen using the feature selection algorithm RFE-SVM and the evaluation of the importance of each feature in the model was done using clustered permutation. **B.** Best features according to the machine learning model to identify the positive seroconversion against COVID-19 after the third dose of the vaccine. The best features were chosen using the feature selection algorithm RFE-SVM and the evaluation of the importance of each feature in the model was done using clustered permutation. **C.** ROC curve of the machine learning algorithm developed for predicting “detectable” after dose 2, as evaluated by nested cross-validation. Curve is mean performance in test groups during 10-fold cross-validation, with fill area the standard deviation. **D.** ROC curve of the machine learning algorithm developed for predicting “positive” after dose 3, as evaluated by nested cross-validation. Curve is mean performance in test groups during 10-fold cross-validation, with fill area the standard deviation.

**Table 3.** Performance of different machine algorithms for vaccine responses in SOT recipients. LR: logistic regression, SVM: support vector machine, RF: random forest, GB: gradient boosting.

| Model | Accuracy | Accuracy (SD) | Balanced Accuracy | Balanced Accuracy (SD) | AUC | AUC (SD) |
| --- | --- | --- | --- | --- | --- | --- |
| **Day 2 (detectable vs undetectable)** |  |  |  |  |  |  |
| LR | 0.824 | 0.101 | 0.825 | 0.083 | 0.900 | 0.095 |
| SVM | 0.917 | 0.093 | 0.921 | 0.084 | 0.950 | 0.081 |
| RF | 0.606 | 0.203 | 0.583 | 0.200 | 0.694 | 0.219 |
| GB | 0.700 | 0.113 | 0.683 | 0.115 | 0.756 | 0.143 |
| **Day 3 (detectable vs undetectable)** |  |  |  |  |  |  |
| LR | 0.598 | 0.167 | 0.596 | 0.168 | 0.560 | 0.242 |
| SVM | 0.814 | 0.131 | 0.813 | 0.137 | 0.854 | 0.179 |
| RF | 0.573 | 0.151 | 0.579 | 0.151 | 0.602 | 0.192 |
| GB | 0.613 | 0.151 | 0.608 | 0.152 | 0.656 | 0.192 |
| **Day 3 (detectable vs undetectable)** |  |  |  |  |  |  |
| LR | 0.714 | 0.162 | 0.730 | 0.173 | 0.845 | 0.129 |
| SVM | 0.795 | 0.133 | 0.802 | 0.161 | 0.933 | 0.118 |
| RF | 0.773 | 0.127 | 0.656 | 0.173 | 0.837 | 0.156 |
| GB | 0.771 | 0.058 | 0.647 | 0.129 | 0.842 | 0.208 |

## Discussion

Impaired immune responses to COVID-19 mRNA vaccination have been well described in SOT recipients.^21–23,41^ However, several studies have shown an improved immune response to mRNA vaccination in these immunocompromised populations after additional boosters^42–44^, highlighting the importance of additional doses of vaccine in these immunocompromised patients to increase their protection against SARS-CoV-2 infection. Whether baseline immune configuration can predict the humoral response to COVID-19 mRNA vaccine among SOT recipients has remained largely unexplored. In our study, SOT recipients showed two distinct immune configuration profiles at baseline. On the one hand, we observed a non-classical immune profile in specific immune parameters that allowed to reach a detectable response after 2 doses of mRNA vaccination, and on the other hand, another immune profile, more similar to healthy adults, allowing to reach a higher antibody response after an additional booster.

Our SOT cohort showed a low rate of seroconversion after 2 doses of mRNA vaccine, increasing after an additional booster (6.6%, 39.8% and 49.4% after 1, 2 and 3 doses of vaccine, respectively). Among SOT recipients, kidney and thoracic transplant recipients have been shown to respond less to COVID-19 vaccine.^45^ Although kidney transplant recipients were more likely to mount a vaccine response above the positive threshold after the third dose in our study, we grouped together both SOT cohorts in order to achieve patient numbers sufficient for downstream statistical analysis. Beyond the low humoral response induced by the vaccination, we wanted to investigate the impact of demographic and clinical variables on vaccine response. Men showed a higher positive vaccine response after dose 3, although not significantly, as Masset *et al.* showed among 136 kidney transplant recipients^46^, whereas antibody responses to bacterial and viral vaccines have been shown to be generally higher in women.^47^ Consistent with a known association of age with reduced COVID-19 vaccine-induced immune responses in the general population^36,48^ and in SOT populations^49^, SOT recipients aged between 50 and 70 years old showed lower immune responses than SOT recipients younger than 50 years old. However, this association was dampened in SOT recipients aged over 70 years old probably in relation to a higher proportion of old grafts (>10 years post-transplant). Indeed, poor vaccine-induced immune response has been observed in recent transplant recipients, particularly in the context of the use of anti-thymocyte globulin (ATG) as induction treatment. This was associated with accelerated immune senescence^50^ in several SOT cohorts^49,51^, highlighting the profound immunosuppression induced by the induction treatment.^52^ In addition, this group of patients aged over 70 years old took a lighter immunosuppressive regimen associated with a better immune response to vaccination in SOT recipients.^37^ Moreover, an eGFR <30 mL/min/1.73m^2^ was negatively associated with the immune response in SOT recipients in our cohort. Uremia has been shown to be associated with a state of immune dysfunction caused on the one hand by the accumulation of pro-inflammatory cytokines due to decreased renal elimination and increased production as a result of uremic toxins, oxidative stress, volume overload, and comorbidities^53,54^ and, on the other hand, by the impact of the uremic milieu and a variety of associated disorders exerted on immunocompetent cells.^38^ According to previous publications^37,45,55,56^, a high number of immunosuppressive treatments and MMF were associated with a lower vaccine-induced immune response. MMF inhibits purine synthesis by reversible inhibition of inosine monophosphate dehydrogenase, in particular the type II enzyme which is expressed predominantly in activated lymphocytes^57^ and has become widely used in SOT populations to prevent allograft rejection in addition to other immunosuppressive treatments, improving the efficacy profiles of immunodepression counterbalanced with detrimental toxicities.^58,59^ Given this profound immunosuppression and the subsequent lower vaccine response in SOT recipients, alternative strategies such as a temporary discontinuation of MMF have been explored in SOT cohorts to increase the immunogenicity of COVID-19 vaccines but revealed controversial results.^60–62^

Regarding specifically the immune cell subsets involved in predictive vaccine response, a few interesting observations can be reported. Focusing on B cells, while total B cell numbers are generally decreased in SOT recipients^63,64^, it is interesting to note that most of the baseline parameters predictive for antibody response either after dose 2 or 3 rely on activation markers (CD80 and CD40) on double negative (DN, i.e. CD27^-^IgD^-^) B cell subsets. DN B cells are mainly increased in the context of aging, infectious or autoimmune diseases^65^, where they have been directly implicated in auto-antibodies secretion in systemic lupus erythematosus.^39^ More importantly these “unconventional B cells” have been involved in influenza vaccine response^40^ and their expansion has been directly linked to protective immunity after influenza vaccine in a younger cohort in comparison to an elderly cohort.^66^ While increased DN B cells have been reported in other SOT cohorts^64^, we mainly observed an increase in DN B cells at baseline in patients responsive after both dose 2 and 3 of COVID-19 vaccine in our cohort (**Supplementary Figure 6A**). Moreover, activation markers such as CD86 have been reported to be decreased in SOT recipients^63,64^ and it is therefore interesting to notice the best vaccine response in patients expressing a higher level of activation markers (CD80 or CD40) on their B cells. Interestingly, total transitional B cells (CD19+CD27-CD38++CD24++) were severely decreased in all our SOT recipients in comparison to healthy controls (**Supplementary Figure 6B**) as previously reported in other studies.^63,64^ Then, we could not confirm in our study that higher baseline levels of transitional B cells could predict antibody response in kidney transplant recipients after COVID-19 vaccine, as reported previously by *Schuller et al.*^64^ and *Stai et al*.^67^

Baseline innate immune cell signature has been linked to protective antibody response after vaccination in various studies.^29,68^ In particular, a pro-inflammatory signature is associated with higher antibody titers in young healthy adults, while the same inflammatory profile has been associated with a worse response in older adults.^29,68^ In our cohort, non-classical monocytes and plasmacytoid dendritic cells were associated with a positive antibody response after the third vaccine dose. As non-classical monocytes are mainly described as anti-inflammatory cells^69^, there is a possibility that our SOT recipients cohort behaves as older adults with a pro-inflammatory myeloid signature being detrimental to antibody response. However, further studies are required to strengthen this hypothesis.

A recent meta-analysis combining transcriptomic data from 13 different vaccines from 28 studies has identified a common baseline immune profile predicting protective antibody titers across various vaccines in healthy controls.^29^ There, baseline immune signature represented by higher abundance of adaptive cell subsets (as opposed to innate pro-inflammatory markers) were generally associated with a lower B cell response on day 7 and overall lower antibody protection after vaccination. Interestingly, in our dataset, similar observations were made with most of the markers associated with low response after the third vaccine dose. These markers were related to different T cell subsets.

Using a machine learning approach to investigate our results, there were several baseline parameters within the NK cell subsets that could contribute to prediction of vaccine response both after the second and third dose. Little is known about the role of NK cells during a vaccine response. They could contribute to the early immune system activation with their innate function through cytokine secretion, as observed during the response after vaccination against Ebola.^70^ Additional roles with adaptive memory function have been attributed to NK cells during specific responses after vaccination against HIV or HBV.^71,72^ Specific subsets such as CD56^dim^CD57^+^ cytotoxic NK cells are mediating this response and similar subsets (CD56^dim^NKG2C^+^) at baseline have been recently identified to predict antibody response after COVID-19 vaccine in healthy adults.^73^ In our cohort, higher baseline activation makers such as CD57, CD94 or CD38 seem to be predictive for a higher antibody level. However, further focused investigation of the role of NK cells in vaccination is required as NK cells can play a double role with regards to their suppression of activated CD8^+^ and CD4^+^ T cells through their cytotoxic activity during vaccine immune responses, as they negatively correlate with protection after adjuvanted HBV or malaria vaccination.^74,75^

While this study identifies different immune correlates associated with detectable or positive responses to the mRNA COVID-19 vaccine Comirnaty in 90 SOT recipients, allowing for in-depth statistical analysis, there are a few limitations. First, the cohort size is limited overall and especially with regards to the number of patients with high antibody levels after vaccination. Additionally, as the immune landscape in SOT recipients is highly influenced by immunosuppressive treatments received during and after transplantation, and are selected based on patient-specific criteria, our cohort presents as a heterogeneous group of patients. The frequent simultaneous administration of different treatments makes it difficult to dissect the effect of single treatments. While several studies have revealed that virus- or vaccine-specific T cells can be present in seronegative healthy individuals or in the context of B cell depletion, for example under treatment for multiple sclerosis^76–78^, we focused on characterizing positive vaccine outcomes based on the humoral virus-specific antibody response. We therefore may classify SOT recipients as non-responders based on their absent seroconversion but do not consider the possibility of successful generation of antigen-specific memory T cells. Moreover, as the immunophenotyping mainly focused on characterization of B and myeloid cell subsets, it did not allow us to perform in-depth characterization of the T cell signature.

An unexpected outcome from this study was the identification of multiple functional immune configurations present in SOT recipients. An immune configuration can, in this case, be defined as a shared meta-phenotype, where individuals share multiple small modifications in immune parameters, which translate into potentially large differences in functional outcomes.^79^ Patients with organ transplantation have undergone multiple severe immunological challenges – the potential initiating pathology, the transplantation with induction treatment, the acute and chronic immune reactions against the graft, and the continued use of immunosuppressive drugs. With the poor response to vaccination exhibited in these patients, a simple model may postulate a single gradient of immune profile, ranging from dysfunctional through to the fully recovered and functional immune state shared with healthy individuals. This model can be supported, to some extent, by the data shown after dose 3: we identified patients with no effective response, and a deviant immune configuration, as well as patients with an effective response, and an immune configuration closer to that of healthy controls. However, unexpected in this model is that we identified another immune configuration that is shared by some SOT recipients capable of detectable immune response after 2 vaccine doses, although for some of them failing to significantly increase their humoral immune response after an additional booster. This immune configuration does not fit within a simple linear immune configuration model, instead achieving a non-classical immune state, highly deviant from the healthy controls, which is, nonetheless, capable of some response to mRNA vaccination. It is tempting to speculate that the non-classical state of this semi-functional configuration allows for a detectable humoral response (Multivariate logistic regression parameters for dose 2 comparison dominated by B cells, including DN B cells), while an additional immune configuration is required for successful seroconversion (with low T cell activation imprint). Further systems vaccinology studies in SOT recipients would be highly informative to verify these observations. Not only could they provide potential diagnostic value for the patients, but knowledge on how these immune configurations are shaped, and their stability over time, may provide broader understanding of how optimal immune responses to vaccination are achieved.

## Methods

### Study design and patient’s characteristics

The study was designed to assess clinical and baseline immunological parameters predicting the humoral response to BioNTech/Pfizer BNT162b2 mRNA (Comirnaty®) vaccination in lung transplant and kidney transplant recipients, naive for SARS-CoV-2 infection. A cohort of health- care workers from Hôpital Saint-Pierre in Brussels was used as a control group regarding baseline immune parameters.

The ethics committee of Hôpital Erasme, Brussels, Belgium (references P2021/182 and A2021/156 for lung transplant recipients; P2020/284 and A2021/131 for kidney transplant recipients) and the Belgian Federal Agency for Medicines and Health Products (FAMHP, EudraCT 2021-004992-16 for lung transplant recipients; EudraCT 2021-000-412-28 for kidney transplant recipients) approved the monocentric prospective phase IV investigator-initiated study of the immunogenicity of the BNT161b2 vaccine (Pfizer-BioNTech). The ethics committee of CHU Saint-Pierre approved a 6-month study on the carriage and seroprevalence of SARS-CoV- 2 in health-care workers (CE/20-04-17).^80^

All participants of at least 18 years of age provided written informed consent for each study, respectively. Lung transplant and kidney transplant recipients from Chest department and Nephrology, dialysis, and transplantation department in Hôpital Erasme, Belgium, respectively and health-care workers from Hôpital Saint-Pierre were enrolled before administration of the first dose of BNT162b2 vaccine. Participants with a previous laboratory-confirmed SARS-CoV-2 infection or with a baseline anti-RBD IgG titer >5 BAU/mL were considered previously infected and excluded from this study. Demographics of the healthy and patient populations are given in **Table 1**. SOT recipients were vaccinated with the Pfizer-BioNTech COVID-19 Vaccine Comirnaty on the day of blood sampling, with second and third doses given on days 21 and between day 150 and 170, respectively. Participants had blood drawn to collect peripheral blood mononuclear cells (PBMCs) just prior to the first vaccine dose (baseline), and serum three weeks after D1 prior to the second dose (post-D1), four weeks after the second dose (post-D2) and four weeks after the third dose (post-D3).

### Sample processing and isolation of PBMCs

Serum tubes were centrifuged at 3000 rpm for 10 min to separate serum. Serum was stored at - 80°C for antibody analysis. PBMCs were isolated from heparinized whole blood by density gradient centrifugation as recommended (Leucosep tubes, Greiner). Whole blood was diluted with an equal amount of Dulbecco’s Phosphate-Buffered Saline with 2% Fetal Bovine Serum and centrifuged. Upper plasma layer was discarded without disturbing the interface. The PBMC layer was retained and washed three times with medium. Cell pellets were diluted with HBSS and aliquoted. PBMCs were frozen progressively with 10% DMSO in FBS and stored in liquid nitrogen.

### SARS-CoV-2-specific binding antibodies by enzyme-linked immunosorbent assay

Binding antibodies at baseline and after vaccination were assessed manually using an enzyme-linked immunosorbent assay (ELISA) for the quantitative detection of IgG-class antibodies to RBD (Receptor Binding Domain, Wuhan strain) [Wantai SARS-CoV-2 IgG ELISA (Quantitative); CE-marked; WS-1396; Beijing Wantai Biological Pharmacy Enterprise Co., Ltd, China] as previously described.^81,82^ For quantification of antibodies, diluted serum samples (1/10, 1/100, 1/400, 1/1600 and 1/6400) were tested with an internal standard, calibrated against NIBSC 20/136 (First WHO International Standard Anti-SARS-CoV-2 Immunoglobulin), and an external positive control sample included on each plate. Diluted samples were incubated (37°C, 30 min) with pre-coated microwells and washed five times. Next, plates were incubated with horseradish peroxidase (HRP)-conjugated anti-human IgG antibodies (37°C, 30 min) and washed five times before adding a TMB and urea peroxide solution for 15 min (37°C°, dark). After incubation, a stop solution (0.5 M H2SO4) was added, and optical density (OD) was measured at 450nm using a Bio-Rad iMark Microplate Absorbance reader (cat#1681130). Net OD values were converted to arbitrary IgG units per mL by interpolation from a point-by-point plot fitted with the standard concentrations and net OD values (correlation coefficient R2≥0.9801), using GraphPad Prism version 9.0.0 for Windows (GraphPad Software, San Diego, California, USA) and exported to Microsoft Excel. Antibody measurements were adjusted for any sample dilution, converted to binding antibody units per mL (BAU/mL) and reported as such. Lower limit of quantification (LLQ) was 5 BAU/mL according to the WHO standard. Clinical performance characteristics of the assay, evaluated in 69 PCR-confirmed COVID-19 patients (comprising mild and severe clinical outcomes, ≥ 15 days post onset of symptoms) and 167 pre-pandemic sera, resulted in a specificity of 100% (95% CI 97,75-100) at a sensitivity of 100% (95% CI 94,73-100) for a cut-off of 6 BAU/mL. Data lower than the limit of detection were attributed the value 2.5 BAU/mL before transformation. Responses reaching the threshold of >264 BAU/mL, as previously used^32,34^, were considered “positive”, while responses >5 BAU/mL were “detectable”.

### Flow cytometry

Frozen PBMCs were thawed in complete RPMI, washed twice with PBS, and stained with live/dead marker (fixable viability dye eFluor780; eBioscience) as well as fluorochrome-conjugated antibodies against surface markers: anti-CD3 (REA613, Miltenyi Biotec), anti-CD123 (7G3), anti-CD80 (L307.4), anti-CD21 (B-ly4), anti-CD27 (L128), anti-BAFF-R (11C1), anti-CD94 (HP-3D9), anti-CD8 (SK1), anti-CD86 (2331 (FUN-1)), anti-CD141 (1A4), anti-CD56 (NCAM16.2), anti-CD4 (SK3), anti-CD16 (3G8), anti-CD40 (5C3, all BD Biosciences), anti-CD57 (HNK-1), anti-CD24 (ML5), anti-HLA-DR (L243), anti-CD19 (HIB19), anti-IgM (MHM-88), anti-CD11c (3.9), anti-IgD (IA6-2), anti-CD10 (HI10a), anti-CD38 (HB-7, all BioLegend), anti-CD14 (TuK4, eBioscience). Cells were then fixed using the Foxp3/Transcription factor buffer staining set (eBioscience) and data were collected on a BD FACSymphony A3 Cell Analyzer (BD Biosciences). A maximum of 5×10^5^ events were acquired for each sample. The data of 5 kidney transplant recipients were excluded from downstream analysis due to insufficient sample quality.

A subset of samples was also used to perform an additional staining using markers focused on the T cell compartment. Cells were thawed and plated as described above, before staining with a live/dead marker (fixable viability dye eFluor780, eBioscience) was performed. After washing, cells were stained with fluorochrome-conjugated antibodies against surface markers: CD14 (61D3), CD4 (RPA-T4), CD45RA (HI100), CCR7 (3D12), CXCR5 (MU5UBEE), ICOS (ISA-3, all eBioscience), CD8 (RPA-T8, Biolegend). Cells were washed again and fixed using the Foxp3/Transcription factor buffer staining set (eBioscience) before acquisition on a BD Canto II (BD Biosciences). A maximum of 2.5x10^5^ events were acquired for each sample. Raw .fcs data were compensated using AutoSpill.^83^ The complete set of FCS files used has been deposited on FlowRepository and annotated in accordance with the MIFlowCyt standard. These files may be downloaded for further analysis from http://flowrepository.org/id/FR-FCM-Z662.

### Flow cytometry gating

Compensated .fcs files were manually pre-processed in FlowJo (LLC, Ashland, Oregon; v10.8) to only contain events with stable fluorescent signals. For samples acquired on the FACSymphony, gating on singlets, leukocytes, live cells, and subpopulations was performed in R (version 4.0.2) by concatenating 7 individual samples and automatically calculating adequate boundaries between populations using the gating strategy outlined in **Supplementary Spreadsheet 1**. These boundaries were then applied to every individual sample to determine cell frequencies and median fluorescence intensity (MFI) values of activation markers on relevant populations (Neumann *et al*., manuscript in preparation). For samples acquired on the BD Canto II, manual gating was performed selecting singlets, lymphocytes and living cells before excluding CD14^+^ monocytes. CD4^+^ and CD8^+^ T cells were further divided based on their CCR7/CD45RA expression profile and expression of ICOS and CXCR5 was evaluated on non- naive cells (excluding CCR7^+^CD45RA^+^ cells). For the gating strategy see **Supplementary Figure 1**.

### Association between demographic and medical variables with vaccine response

To assess the association of demographic characteristics, comorbidities and treatments with the vaccine response induced by mRNA vaccination, sex and age were considered *a priori* confounders, i.e., always considered confounders regardless of their effect on the relationship. The other demographic variables and treatment regimens were considered potential confounders. They were only considered confounding if a significant impact on the model between exposure evaluated and the outcome was observed. The relationship was evaluated using multivariate logistic regression. Stepwise regression bidirectional elimination was used to select among potential confounders. The stepwise criterion of permanence was a significant p-value (<0.05) of the potential confounder, thus characterizing its importance for the model.

### Batch correction

For flow cytometry, a batch variation in channel intensity is inherent to the measurement process. Here, we first verified whether there were batch differences in the cytometry data. We found flow cytometry readouts with a significant difference between batches (t Student/Mann-Whitney test), while other demographic/treatment variables of the study did not show significant differences between batches (t Student/Mann-Whitney test for continuous variable and Pearson’s chi-squared for categorical variable). This indicated that the batch effect differences were due to technical variation in the flow cytometer and staining procedure. To correct measurement distortions due to day-to-day variation we located a cell population that had the smallest proportional variation between samples for each immune marker (considering populations of cells with more than 30 cells) and used their mean to correct the respective marker channel.

### Identification of immune variables associated with vaccine response

For each of the 444 immunological variables, we assessed their association with vaccine response by considering only SOT recipients divided based on: i) the presence of antibodies against SARS-CoV-2 detectable/undetectable after the second dose; ii) the presence of antibodies against SARS-CoV-2 detectable/undetectable after the third dose; and iii) the presence of antibodies against SARS-CoV-2 above the positive threshold/below the positive threshold after the third dose. This stratification of patients was based on the very low number of SOT recipients reaching the positive threshold, particularly after only two doses, requiring analysis to focus on achieving the detectable antibody reading, or on reaching the positive threshold after the third dose only. As each immune marker may have different levels of measurements, to compare the magnitude of the effect of each marker on the vaccine response, a z-score transformation was applied. The evaluations of the associations of the immune markers in vaccine response were performed through multivariate logistic regression. Sex and age were considered *a priori* confounders. Stepwise regression bidirectional selection^84^ was used to select additional potential confounders. The stepwise criterion for selecting features was Akaike information criterion^85^, keeping only potential confounders that improve the model. This resulted in adjustment for sex, age, type of transplant, and the time since transplant. Among the immunological markers that obtained p-values lower than 0.05, the 10 that had the greatest magnitudes of effect were chosen for further analysis. Logistic regression models were run in Python using the package statsmodels version 0.13.2.

### Similarities between immune markers in healthy and transplant recipients

To understand the similarity of the immune response between healthy controls and SOT recipients, UMAP dimension reduction technique^86^ was performed. To do so, markers with more than 30% of missing values were first removed and subsequently the remaining missing values were imputed using the mean. After that, a standardization transformation was performed. To assess the distances between individuals, the Calinski and Harabasz score^87^ was used. In addition to understanding the immune similarity between SOT recipients and healthy controls, it is also important to understand whether the relationships between immune markers change within individuals. For that we used a heatmap for Spearman correlation and compared these correlations in SOT recipients with healthy controls.

### Machine learning

Data was cleaned and standardized as described above with standardization applied within each group. For selection of important markers contributing to the outcome by machine learning, we used recursive feature elimination^88^, with 10-fold cross-validation using the area under Receiver Operating Characteristic (ROC) curve as readout. The model used for feature selection was support vector machine (SVM). The choice of model hyperparameters was performed by grid search with 3-fold cross validation using balanced accuracy as a selection criterion.

Due to the multicollinearity shared between the markers, the potential importance of the marker can be underestimated as there are other similar markers contributing similar information. Therefore, to limit this potential loss of important markers, we applied hierarchical clustering of markers based on the dissociation of the markers given 1 minus modulus of the Spearman correlation. After the identification of these clusters, the assessment of the importance of each marker was performed using permutation importance^89^ using all markers and excluding markers contained in the same group of the evaluated marker. Thus, the marker’s importance in the phenotype is its impact on the model containing only itself in its group and a marker from every other group. The model used in the permutation presented the same hyperparameters selected in the feature selection. Permutation was performed with 100 repetitions and evaluated by area under ROC curve (AUC).

The machine learning models were created using the best features to reduce dimensionality. The best models were selected and evaluated by nested cross-validation, applying 10 external and 5 internal cross-validations. The selection of the hyperparameters of the models was done by grid search using balanced accuracy as a criterion.

### Data and code availability

Raw data for the study is available from https://flowrepository.org/experiments/6338 (FR-FCM- Z662). Source data for the analysis presented in the main and supplementary text is available as **Supplementary Spreadsheet 2**. Source code for the analysis is available on GitHub at: https://github.com/rafael-veiga/immunological_covid.

## Data Availability

All data produced are available online at http://flowrepository.org/id/FR-FCM-Z662

## Acknowledgments

The authors thank all patients and volunteers enrolled in the study. This study was co-funded by the Belgian Federal Government through Sciensano and by the Fonds de la Recherche Scientifique, F.R.S.-FNRS, Belgium. This work has received funding from the KU Leuven C1 program, the European Union’s Horizon 2020 research and innovation programme under grant agreement No 874707 (EXIMIOUS) (to SHB and AL), and the Biotechnology and Biological Sciences Research Council (BBSRC) through Institute Strategic Program Grant funding BBS/E/B/000C0427 and BBS/E/B/000C0428 and the KU Leuven BOFZAP start-up grant (to SHB). This study was supported by the European Regional Development Fund (ERDF) of the Walloon Region (Wallonia-Biomed portfolio, 411132-957270). The authors acknowledge the important contributions of Jeason Haughton, Reena Chinnaraj and the KU Leuven FACS Core. NG received a PhD studentship from the Fonds Erasme, Belgium and from the F.R.S- FNRS, Belgium. JN is an FWO fellow. DK received a PhD studentship from the F.R.S- FNRS, Belgium. MG is supported by the Belgian Kids fund (BKF). N.D. is a clinical researcher of the F.R.S.- FNRS. SG and AM are research directors of the FRS-FNRS. SHB is supported by VLAIO (Flanders Innovation & Entrepreneurship) within the project entitled PRISMA and by a grant from Stichting Alzheimer Onderzoek - Fondation Recherche Maladie Alzheimer (SAO-FMA) and Stichting tegen Kanker (grant: 2020-106 and 2022-140). The authors declare no conflict of interest.

---

**Supplementary Figure 1.**
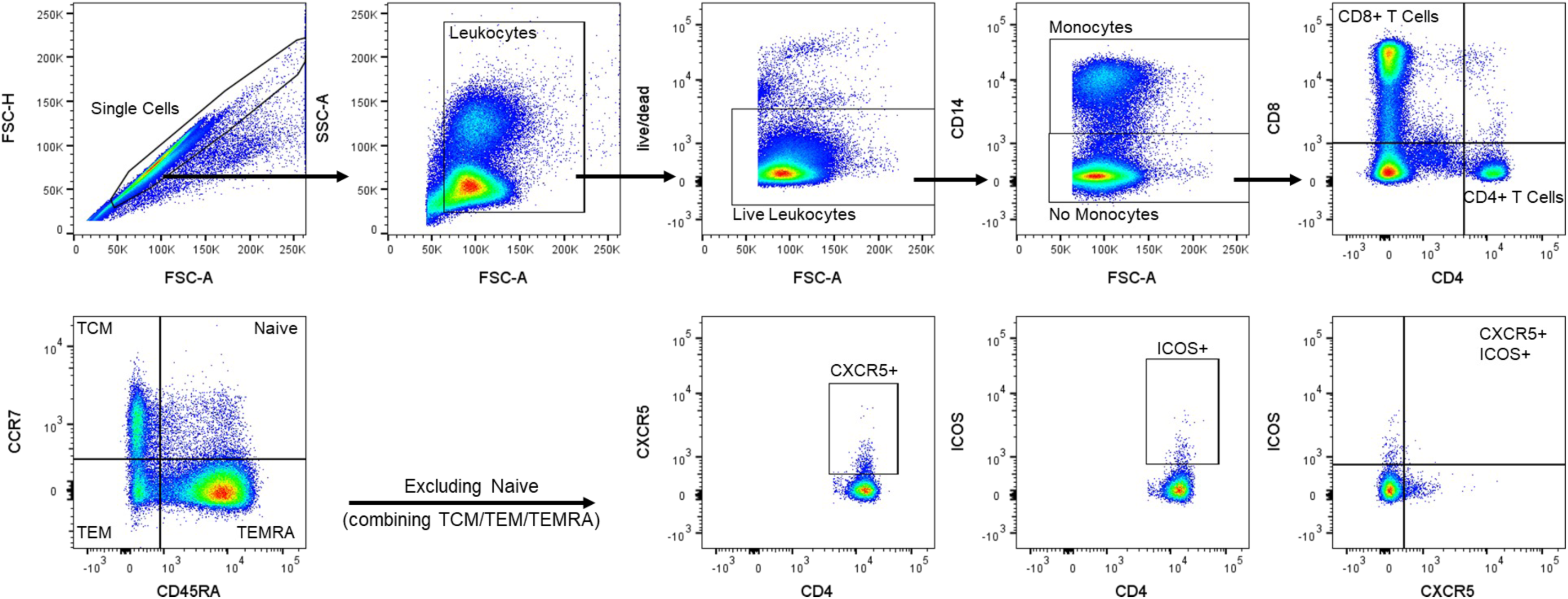
Manual gating strategy for T cell phenotyping. All acquired events were gated on single live cells before excluding CD14^+^ monocytes. CD4^+^ conventional T cells and CD8^+^ cytotoxic T cells were further separated into naive and memory cells based on expression of CCR7 and CD45RA. CXCR5 and ICOS expression was used to identify T follicular helper cells.

**Supplementary Figure 2.**
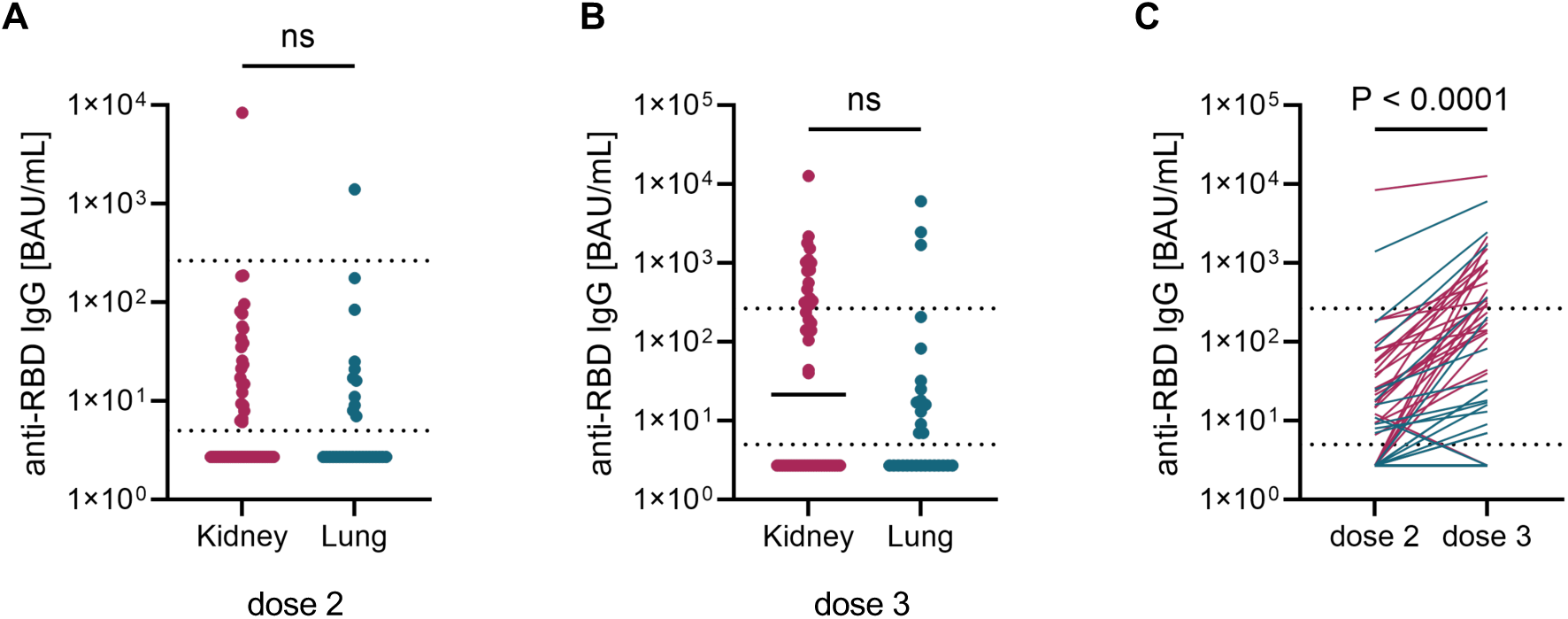
Anti-RBD IgG antibody levels increase after a third vaccine dose in SOT recipients. Levels of SARS-CoV-2-specific anti-receptor binding domain (RBD) IgG antibodies were measured using ELISA following **A.** two or **B.** three doses of mRNA vaccine. **C.** The change in antibody levels is shown for separate individuals with lung recipients indicated in blue and kidney recipients indicated in red. Statistics in **A** and **B** are based on Mann-Whitney-U and in **C** on Wilcoxon rank comparisons. Of note, binding antibody levels of 57 kidney transplant recipients are represented after dose 2 because of 2 data not available, binding antibody levels of 50 kidney transplant recipients are represented after dose 3 because of 5 data not available and 4 patients with COVID-19 before dose 3, binding antibody levels of 29 lung transplant recipients are represented after dose 3 because of 2 data not available.

**Supplementary Figure 3.**
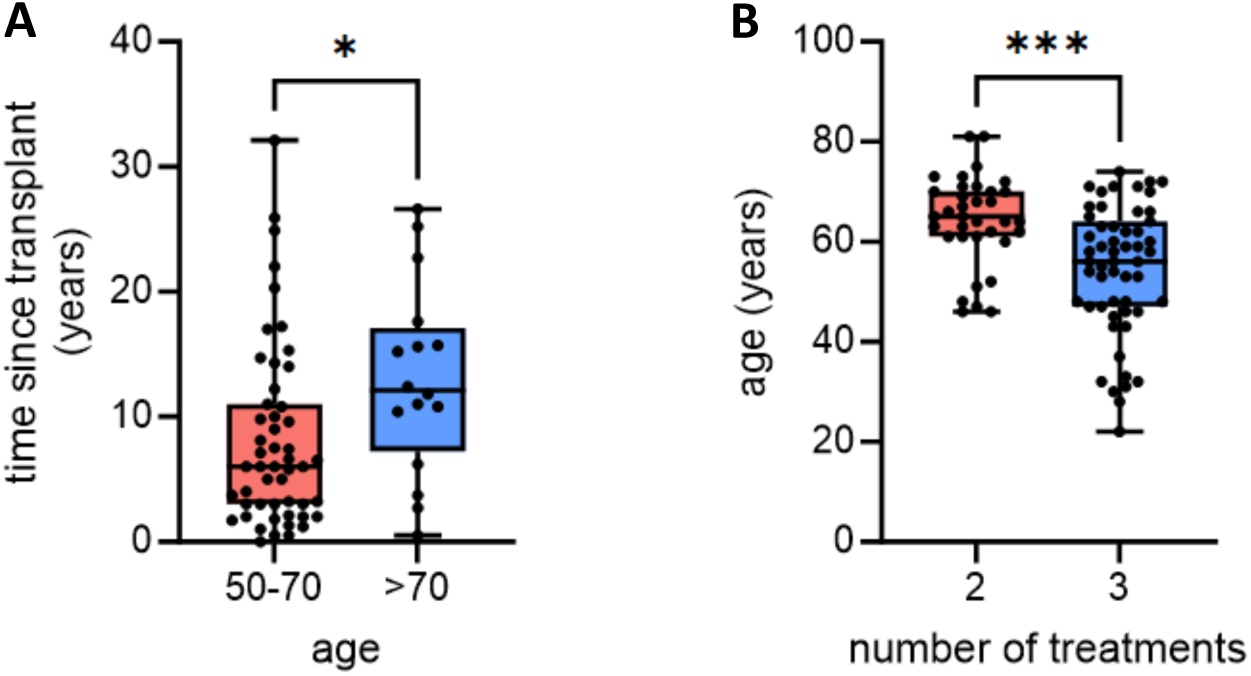
Age-association with time since transplant and combined number of treatments. **A.** Association between time since transplant in years and age, grouped as 50-70 or >70 years. **B.** Age of patients receiving either two or three combined treatments. Statistics are based on Mann-Whitney-U. * = p<0.05; *** = p<0.001.

**Supplementary Figure 4.**
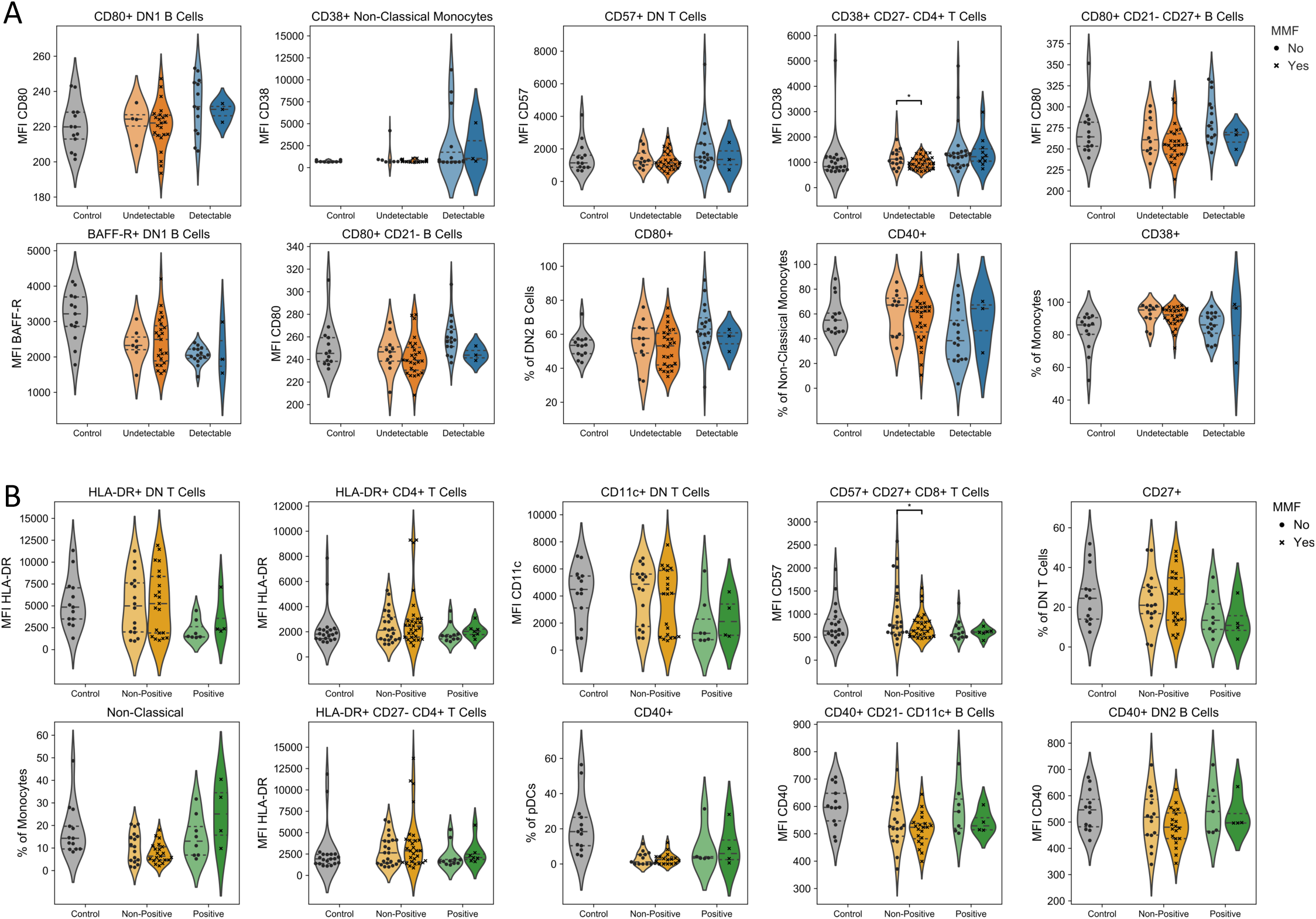
Influence of MMF treatment on key immune parameters associated with vaccine response in SOT recipients. 110 individuals were recruited to a systems vaccinology study, including 20 healthy controls, 31 lung transplant recipients and 59 kidney transplant recipients. Immunological profiles were assessed at baseline (day 0 of vaccination) for 444 immunological variables. Multivariate logistic regression was performed adjusting by type of transplant, sex, age and the number of years since transplantation, identifying the 10 highest magnitude effects (based on log(OR of the z-score)). **A.** Raw value graphs, showing healthy controls, dose 2 non-detectable and dose 2 detectable SOT recipients, for the 10 highest magnitude parameters associated with dose 2 antibody detection stratified by treatment with MMF (circle indicates no MMF treatment, cross indicates MMF treatment). Individual values, no imputation. **B.** Raw value graphs, showing healthy controls, dose 3 non-positive and dose 3 positive SOT recipients, for the 10 highest magnitude parameters associated with dose 3 antibody positive seroconversion stratified by treatment with MMF (circle indicates no MMF treatment, cross indicates MMF treatment). Individual values, no imputation. Statistics are based on two-tailed individual t-tests comparing patients receiving treatment or not within each response group and only significant comparisons are shown. * = p<0.05.

**Supplementary Figure 5.**
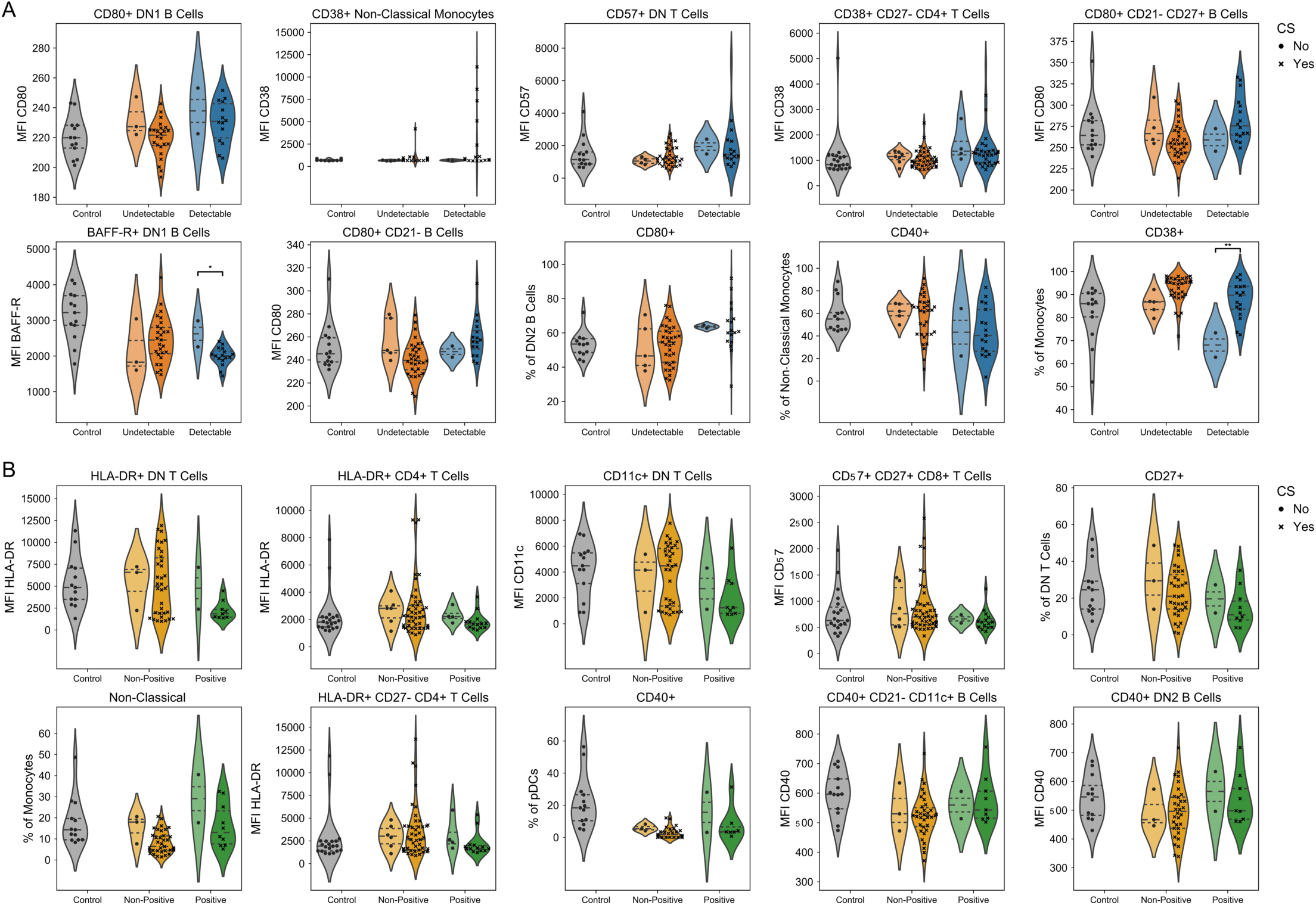
Influence of CS treatment on key immune parameters associated with vaccine response in SOT recipients. 110 individuals were recruited to a systems vaccinology study, including 20 healthy controls, 31 lung transplant recipients and 59 kidney transplant recipients. Immunological profiles were assessed at baseline (day 0 of vaccination) for 444 immunological variables. Multivariate logistic regression was performed adjusting by type of transplant, sex, age and the number of years since transplantation, identifying the 10 highest magnitude effects (based on log(OR of the z-score)). **A.** Raw value graphs, showing healthy controls, dose 2 non-detectable and dose 2 detectable SOT recipients, for the 10 highest magnitude parameters associated with dose 2 antibody detection stratified by treatment with corticosteroids (CS; circle indicates no CS treatment, cross indicates CS treatment). Individual values, no imputation. **B.** Raw value graphs, showing healthy controls, dose 3 non-positive and dose 3 positive SOT recipients, for the 10 highest magnitude parameters associated with dose 3 antibody positive seroconversion stratified by treatment with CS (circle indicates no CS treatment, cross indicates CS treatment). Individual values, no imputation. Statistics are based on two-tailed individual t-tests comparing patients receiving treatment or not within each response group and only significant comparisons are shown. * = p<0.05; ** = p<0.01.

**Supplementary Figure 6.**
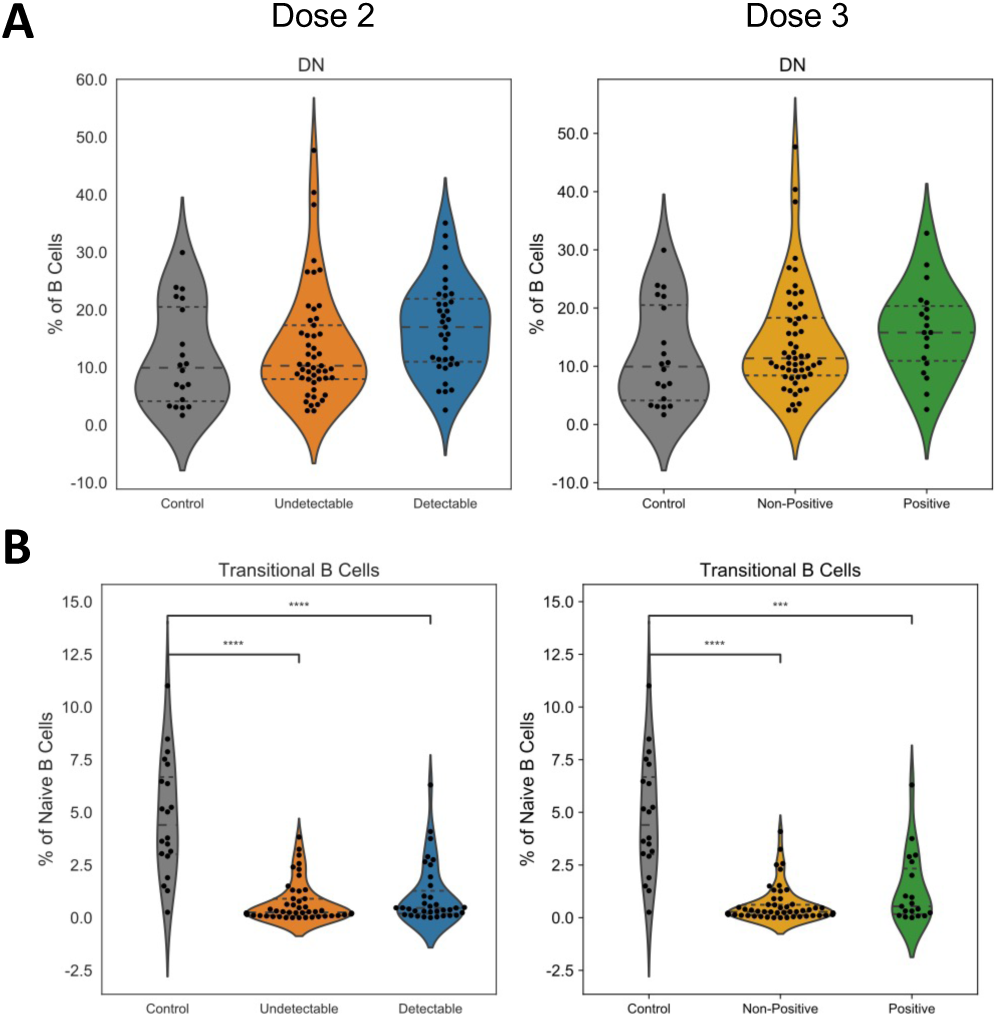
Changes in DN and transitional B cell subsets associated with effective SARS-CoV-2 vaccine response in SOT recipients. 110 individuals were recruited to a systems vaccinology study, including 20 healthy controls, 31 lung transplant recipients and 59 kidney transplant recipients. Immunological profiles were assessed at baseline (day 0 of vaccination) for 444 immunological variables. **A.** Raw value graphs, showing healthy control, dose 2 non-detectable and dose 2 detectable SOT recipients (left panel) and healthy controls, dose 3 non-positive and dose 3 positive SOT recipients (right panel), for double negative (DN) B cells. Individual values, no imputation. **B.** Raw value graphs, showing healthy controls, dose 2 non-detectable and dose 2 detectable SOT recipients (left panel) and healthy controls, dose 3 non-positive and dose 3 positive SOT recipients (right panel), for double negative (DN) B cells. Individual values, no imputation. Statistics are based on Kruskal-Wallis with Dunn’s post-hoc comparison and only significant comparisons are shown. *** = p<0.001; **** = p<0.0001.

